# SPC: a SPectral Component approach leveraging Identity-by-Descent graphs to address recent population structure in genomic analysis

**DOI:** 10.1101/2025.06.04.25328990

**Authors:** Ruhollah Shemirani, Gillian M. Belbin, Sinead Cullina, Christa Caggiano, Christopher Gignoux, Noah Zaitlen, Eimear E. Kenny

## Abstract

Population structure is a well-known confounder in statistical genetics, particularly in genome-wide association studies (GWAS), where it can lead to inflated test statistics and spurious associations. Traditional methods, such as principal components (PCs), commonly used to adjust for population structure, are limited in capturing fine-scale, non-linear patterns that arise from recent demographic events – patterns that are crucial for understanding rare variant effects. To address this challenge, we propose a novel method called SPectral Components (SPCs), which leverages identity-by-descent (IBD) graphs to capture and transform local, non-linear fine-scale population structure into continuous representations that can be seamlessly integrated into genetic analysis pipelines. Using both simulated datasets and empirical data from the UK Biobank (N ≈ 420,000), we demonstrate that SPCs outperform PCs in adjusting for fine-scale population structure. In simulations, SPCs explained over 90% of the fine-scale population structure with fewer components, while PCs captured less than 5%. In the UK Biobank, SPCs reduced the inflation of p-values in the GWAS of an environmental-driven phenotype by 12% compared to PCs, while maintaining a similar performance to PCs in height, a highly heritable phenotype. Additionally, SPCs improved rare variant association analyses, reducing genomic inflation (e.g., from 7.6 to 1.2 in one analysis), and provided more accurate heritability estimates. Spatial autocorrelation analysis further confirmed the ability of SPCs to account for environmental effects, reducing Moran’s I for both environmental and heritable phenotypes more effectively than PCs. Overall, our findings demonstrate that SPCs provide a robust, scalable adjustment for recent population structure, offering a powerful alternative or complement to PCs in large-scale biobank studies.

## Introduction

Population structure is a critical factor in statistical genetic studies, as it can confound results if not properly addressed (Price et al. 2006; Lawson et al. 2020). This structure arises from genetic and environmental differences across populations, shaped by historical processes such as migration (Gravel et al. 2013), genetic drift (Nei and Tajima 1981), selection (Berg et al. 2019; Sohail et al. 2019; Mathieson et al. 2023), and assortative mating (Border et al. 2022). These processes create systematic differences in variant frequencies within and between populations that can violate key statistical assumptions in genetic analyses (Sohail et al. 2019; Price et al. 2010). For example, population structure can violate the assumption of independence among individuals in genetic analyses, such as Genome Wide Association Studies (GWAS), leading to false-positive associations and inflated test statistics (Cook et al. 2020). To mitigate these effects, genetic analysis pipelines often use Principal Components (PCs; Price et al. 2006) and/or (generalized) linear mixed models (GLMMs; Kang et al. 2010). PCs represent major axes of genetic variation derived from dimensionality reduction of a genotype matrix, or the Genetic Relatedness Matrix (GRM), which stores pairwise genetic similarity based on variants. GLMMs complement this approach by simultaneously accounting for population structure and cryptic relatedness by modeling a random effect whose covariance structure is defined by the estimated GRM. Both PC- and GLMM-based strategies have been successfully used to address population structure in analysis involving common variants. However, it is less clear whether these methods will be similarly effective for rare variants and more complex fine-scale population structure (Mathieson and McVean 2012; Zaidi and Mathieson 2020; Privé et al. 2020).

Large-scale sequencing studies have revealed patterns of recently arisen rare variants and have shown that they tend to be geographically localized (Fu et al. 2013; Gravel et al. 2011). This fine-scale population structure impacting rare variants has been detected in several populations, including Icelandic (Helgason et al. 2005), British (Cook et al. 2020; Gilbert et al. 2022; Nait Saada et al. 2020), and Japanese (Sakaue et al. 2020), revealing distinct genetic patterns shaped by historical movements over the past few thousand years. However, these studies also highlight a critical challenge: traditional methods to account for population structure struggle to capture this discrete, non-linear population structure occurring within recent timescales. The non-linear relationships and sparsity in the GRM introduced by rare variants can make rare variant PCs sensitive to noise and violate the linear assumptions of PCA, complicating the interpretation of results (Baye et al. 2011; Ma and Shi 2020; Zaidi and Mathieson 2020). Likewise, incorporating rare variant information in GLMMs introduces several limitations, including higher computational costs and reduced power compared to common variant counterparts (S. Lee et al. 2014; Wainschtein et al. 2022). These findings underscore the need for novel analytical methods tailored to the unique properties of rare variants in statistical genetic analyses, enabling more accurate modeling of fine-scale population structure and the complex genetic architecture they reveal.

To address these limitations, estimates of pairwise haplotypes shared identical-by-descent, or Identity-By-Descent (IBD), offers a more precise approach for detecting recent fine-scale population structure in large genomic datasets (Shemirani et al. 2021). The length of shared IBD segments provides a direct measure of evolutionary time – longer segments indicate more recent population structure, typically within the past dozen generations (Browning and Browning 2015). At the population level, the aggregated lengths of IBD segments shared between all pairs of individuals across the genome can be utilized to construct graphs of genetic similarity, where individuals are represented as nodes and shared IBD haplotypes are represented as edges (Belbin et al. 2021). Previous studies have shown the advantages of utilizing IBD data to represent recent population structure through the extraction of clusters of individuals who share IBD haplotypes across the genome (Belbin et al. 2021; Caggiano et al. 2023; Dai et al. 2020). These IBD-based clusters reveal population structure missed by traditional methods, leading to differential associations with health outcomes, unique enrichment of casual rare variants, and distinctive performance of polygenic risk scores compared to clusters identified with common variants alone (Belbin et al. 2021; Johnson et al. 2022).

Here, we introduce SPectral Components (SPCs), a graph theory-driven approach that transform the discrete, localized signals of recent population structure, represented by an IBD graph, into a continuous representation of genetic similarity that can be used in statistical genetic analysis pipelines. Our method leverages graph Laplacian transformations (Grone et al. 1990), a fundamental concept in computational fields, such as dimensionality reduction (Kunegis et al. 2010), graph-based machine learning (Si et al. 2014), and network analysis (Shuman et al. 2013), due to their ability to encode graph topology and smooth representations of variations in node features across the graph (Shuman et al. 2013). Previous work has applied spectral approaches to common variant data and demonstrated they can be more robust than PCs in finding hidden population structure compared to GRMs (A. B. Lee et al. 2010). In this study, we use simulated and empirical data from the UK Biobank project to explore the efficacy of SPCs in adjusting for the effects of population structure compared to traditional methods. We also examine the performance of these approaches in rare variant analysis under environmental confounding. We demonstrate how SPCs optimally extract patterns of population structure that are resilient to the presence of discrete and localized structure in both variant and environmental association, via leveraging graph spectral decomposition.

## Results

### Overview of Spectral Components (SPC) Calculation

Previous work has demonstrated the efficacy of identity-by-descent (IBD) graphs in capturing fine-scale population structure signals absent from GRMs (Zaidi and Mathieson 2020; Caggiano et al. 2023; Browning and Browning 2023). We developed a pipeline to extract individuals frequency components underlying a graph of IBD sharing at a population level, or Spectral Components (SPCs; **Figure 1**). SPCs were calculated in 4 steps. First, we phased genotype data, and estimated IBD using iLASH (Shemirani et al. 2021), We then aggregated the total sums of IBD sharing for each pair of individuals, which were then utilized to generate an undirected, unweighted relatedness graph, with each participant represented as a node. IBD relatedness in this graph was represented as connecting edges between respective nodes, if their sum of IBD sharing surpassed a minimum threshold of 6cM. Thirdly, we generated an adjacency matrix representation of this graph. Lastly, we calculated the spectral components of this matrix as SPCs. To conduct comparative analysis, we further calculated alternative principal components of this graph, along with the principal components of the original genotype data, and rare variants.

**Figure 1.**
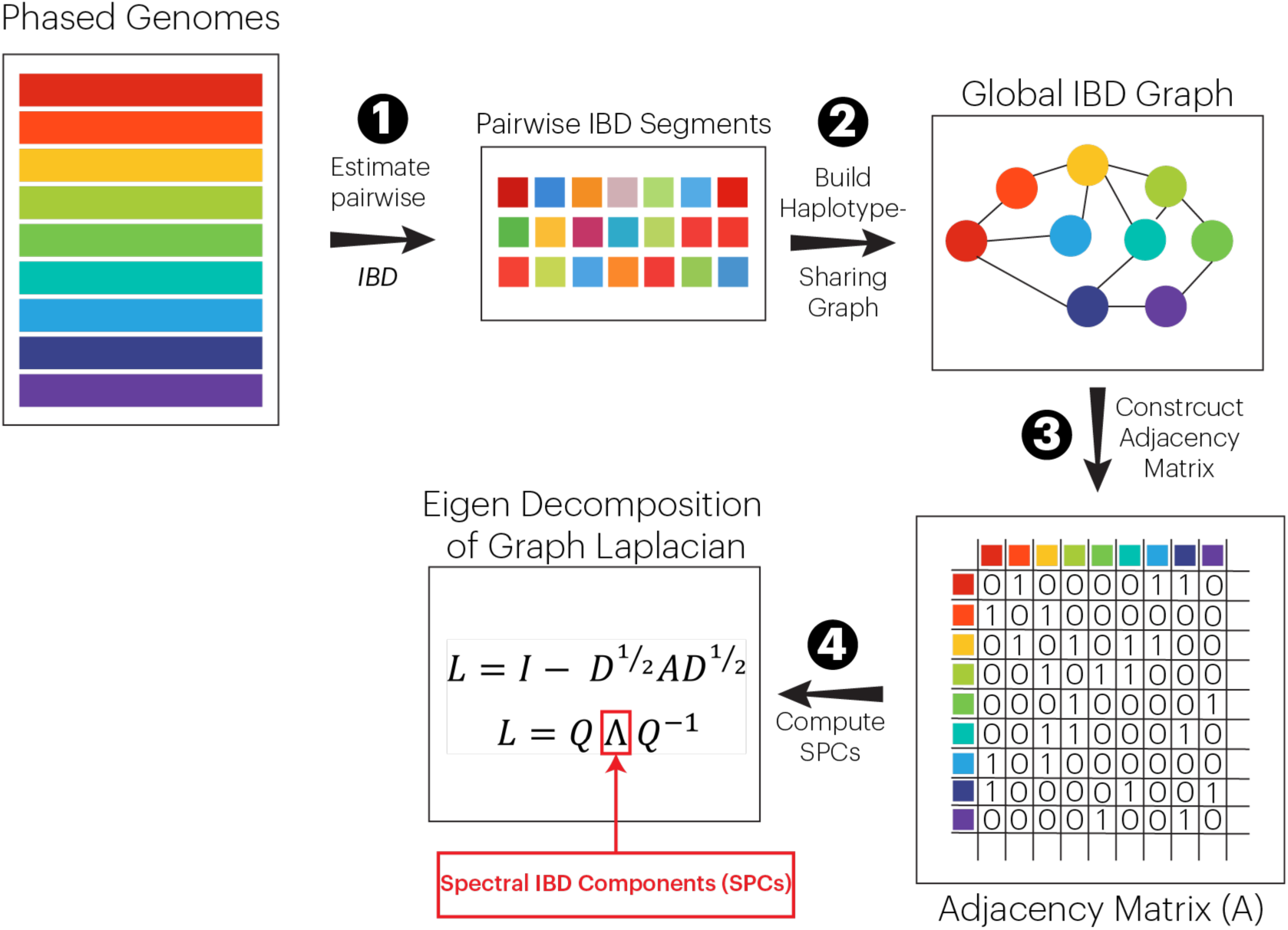
General schema for calculating SPCs. First, phased genotypes are used to estimate segments of genome shared between individuals IBD. Second, the IBD segments are utilized to generate a relatedness graph with vertices representing participants and edges representing aggregated IBD sharing between pairs of participants above a threshold of 6cM genome wide. Third, the IBD relatedness graph is transformed into an adjacency matrix (A). Finally, the adjacency matrix A is transformed into its Laplacian form. The eigenvectors corresponding to the smallest eigenvalues of this matrix comprise the set of SPCs.

### Evaluating Performance to Detect Fine-scale Population Structure on a Simulated Dataset

To evaluate the performance of SPCs for capturing fine-scale population structure in genomic datasets, we first simulated a population, following Zaidi & Mathieson, 2020. We simulated a 5-by-5 grid of 25 demes with a low migration rate among them that emulates a homogeneous population structure (**Figure 2a**). This configuration closely resembles the recent fine-scale population structure observed in the White British cohort in the UK Biobank (see Methods). By simulating this nuanced structure, we aimed to evaluate how well SPCs can discern subtle genetic differences that are often overlooked by traditional methods, thereby providing a more detailed understanding of population stratification.

**Figure 2.**
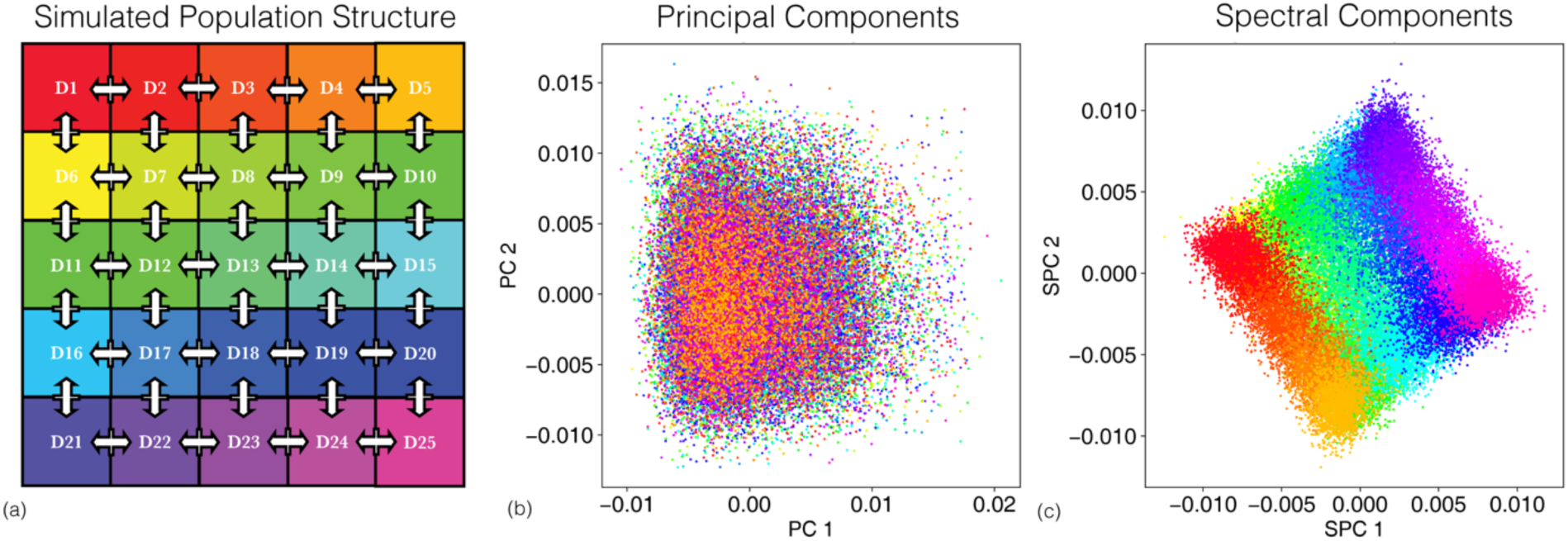
Reconstruction of recent population structure by covariates. **a**) A schematic of our simulation of a cohort with homogeneous population structure with 25 demes and constant migration (5% per generation) between neighboring demes. All demes coalesce to single ancestral deme 200 generations ago. Consequently, simulated individuals have similar genetic backgrounds in terms of common variants, while simultaneously harboring distinct structure in terms of rare variation and environment of origin. **b, c**) PC and SPC representation of the shared genetic ancestry among simulated samples. Dots in either panel represent samples colored based on the deme of origin.

The simulated dataset included 50,000 diploid samples. Only the first chromosome was simulated for computational tractability. We calculated PCs of common and rare variants. We then down sampled the dataset to retain only the common variants, phased the data to replicate phasing artifacts, and estimated aggregated IBD sharing using iLASH.

Compared to PCs, SPCs demonstrate significant improvements in capturing population structure (**Figure 2b-c**). We used correlation with deme of origin as a basic measurement of the ability of SPCs to dissect fine-scale population structure. We indexed demes based on their order on horizontal and vertical axes to generate coordinates of origin for each simulated sample, akin to place of birth for an individual. The first 100 PCs explain 4% and 5% of the variation in the vertical and horizontal coordinates of the deme of origin for simulated samples, respectively. However, the first 10 SPCs were sufficient to explain 90% of the variation in the same categories of data. This is reflected in better visual separation between samples based on their deme of origin.

### Evaluating Performance for Disentangling Environmental and Genetic effects

To evaluate the performance of SPCs as covariates in genomic analysis pipelines, we simulated 7 continuous phenotypes (**Figure 3a**), two of which were non-heritable and 5 with different heritability schemes. The first, referred to as ‘environmental smooth’, was a continuous phenotype in which variation solely originated from indirect environmental effects. The mean value of the phenotype decreased from North to South (**Figure 3a**), and the effects of linear dependencies on deme of origin among the mean values of outcomes were observed. The second, which Zaidi and Mathieson called ‘environmental sharp’(Zaidi and Mathieson 2020), aimed to investigate the effects resulting from the invalidity of common linear assumptions (**Figure 3a**). For this phenotype, a single target deme had a phenotype distribution of 𝑦∼ 𝒩(𝜇, 2𝜎) with a non-zero 𝜇, while other demes had a phenotype distribution of 𝑦∼ 𝒩(0, 𝜎); mimicking non-linear indirect environmental effects. Due to variation stemming only from environmental effects, the expected narrow-sense heritability of both environmental phenotypes simulated is zero.

**Figure 3.**
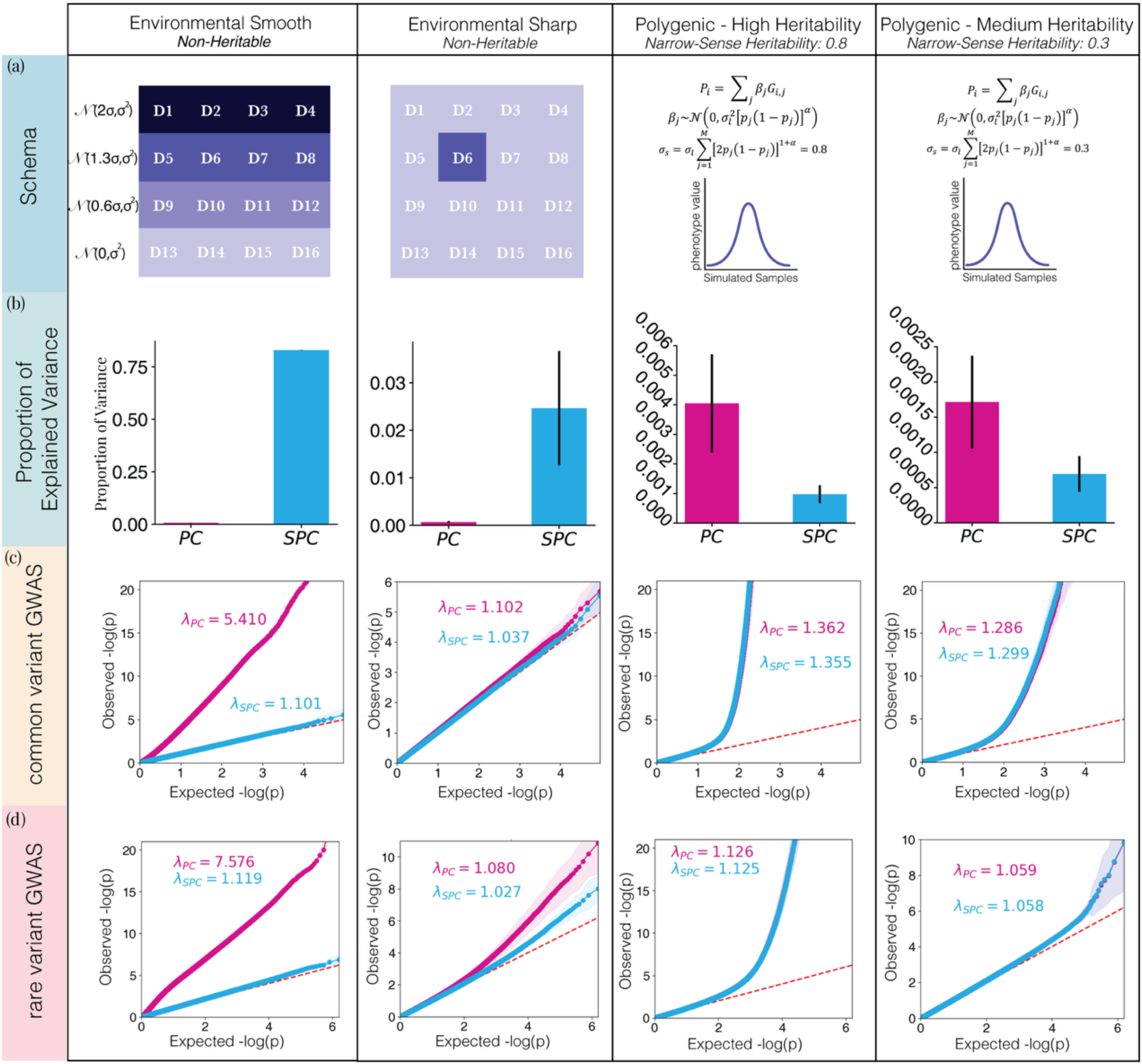
Comparison of PCs and SPCs as covariates to correct for population structure on the outcomes in simulations. **a)** The schema of simulated phenotypes. First, the environmental smooth phenotype, where the values are determined based on the vertical coordinate of origin for each individual. The phenotype values are drawn from a normal distribution with shared standard deviation. The mean of the distribution in each row is lower than the row above, resulting in a difference of twice the standard deviation between the top and the bottom row. Second, the environmental sharp phenotype is determined by the deme of origin. The phenotype values for all demes other than D6 is drawn from a zero-centered normal distribution, whereas in D6, phenotype values are drawn from a normal distribution with the mean set to twice the standard deviation. Finally, the two polygenic phenotypes were determined by assigning effect sizes to causal variants, randomly selected from windows of 10,000 base pairs, and calculating the polygenic score based on the effect sizes. Effect sizes were drawn from a random normal distribution with a mean of zero and a variance derived from minor allele frequency, heritability, and selective pressure. 𝜎_s_ determines the expected heritability of the phenotype, while 𝛼 determines the selective pressure, with negative values resulting in higher effect sizes assigned to variant with low minor allele frequency. Tested values for 𝜎_s_ were 0.8 for the high heritability phenotype and 0.3 for the medium heritability phenotype. 𝛼 was set to −0.5 across both experiments **b)** proportion of variation in the phenotypes explained by the first 25 PCs and SPCs in 4 categories of simulated phenotypes. Adjustment methods are listed on the x-axes, while the y-axes represent proportion of variance explained by each model. The error bars represent the standard deviation. **c)** Inflation of p-values in a GWAS analysis of simulated phenotypes using common variants only. **d)** Inflation of p-values in a GWAS analysis of simulated phenotypes using rare variants (10<MAC<100).

The next 3 phenotypes, on the other hand, were heritable (h^2^=0.1, 0.3, 0.8), and were affected only by direct genetic effects and random noise. These phenotypes, which we refer to as “polygenic phenotypes”, are modeled through the randomized selection of causal variants with effect sizes assigned based on their allele frequency and the total heritability of the trait. This approach of simulating effect sizes results in negligible correlation with the deme of birth, as it does not allocate a high impact to common variants that undergo significant drift, subsequently impacting mean phenotype values per deme (Schoech et al. 2019). Absence of such confounding effects eliminate both the biases described earlier and the requirement to adjust for them. Thus, fixed-effect covariates, such as PCs, are not effective in addressing possible confounding effects for this phenotype. Instead, Linear Mixed Models can be used in such scenarios to account for variance components induced by genetic effects (Price et al. 2010).

The last two phenotypes, henceforth referred to as “hybrid” phenotypes, combine environmental and polygenic phenotypes via replacing the random noise components of the polygenic phenotype with environmental phenotypes. The hybrid smooth phenotype is generated via adding the polygenic phenotype with heritability 0.3 and the environmental smooth phenotype (reweighted to comprise 0.7 of the variance of the phenotype). The hybrid sharp phenotype is generated similarly using the environmental sharp with the same weighting scheme. The environmental category was designed to evaluate Type-1 error rate, while the polygenic phenotypes assess power. Finally, the hybrid phenotypes evaluate how well-calibrated each model is in the presence of both background polygenic effects and confounding environmental effects.

To measure the efficacy of SPCs in adjusting for environmental effects on phenotypes, we used proportion of variation in the phenotype explained by covariates (PVE; **Figure 3b, Supplemental Table 1**). For the smooth phenotype, the first 25 SPCs accounted for 83% of the PVE, while the same number of PCs explained only 0.7%. For the sharp phenotype, the PVE for both models decreased, although the SPCs still explained 40-fold more environmental effects than PCs (2.5% of PVE vs 0.06%). SPCs achieved their lowest PVE in the analysis of the heritable phenotypes with no environmental effects from recent population structure, where both models explained less than 0.5% in the variation of the phenotypes across all heritability models (**Supplemental Figure 1**). PVE values from the PC-based model were slightly higher than the SPC-based models (0.1% vs. 0.4%, respectively). Analyzing the hybrid smooth phenotypes as outcomes resulted in a PVEs of 58% and 0.7% for the SPC-based, and PC-based models respectively. PVEs for the hybrid sharp phenotype were 1.7% and 0.2% for the two models, respectively (**Supplemental Figure 1**). Overall, these simulations demonstrate that SPCs more effectively capture environmental effects compared to PCs, while not significantly impacting estimates of true heritable effects.

### Evaluating Performance for Genome Wide Association Studies

To evaluate how SPCs perform in the context of genome-wide association studies, we conducted GWAS using all 7 phenotypes, focusing on the distribution of z-scores and the inflation of p-values as a measure of the unadjusted effects of population structure (Devlin and Roeder 1999; Yang et al. 2011) (**Figure 3c**). In the GWAS of common variants for the smooth phenotype, the genomic inflation factor (denoted by 𝜆_gc_) was 1.101 [confidence intervals (CI): 1.084-1.118] when including

SPCs in the model, compared to 5.410 [CI: 5.305-5.517] when including PCs in the model, indicating that SPCs were better controlling for population structure. The difference in performance in the sharp model was less pronounced, although statistically significant, 1.102 [CI: 1.066-1.138] for the PC-based models and 1.037 [CI: 1.019-1.054] for the SPC-based ones. This pattern was not observed in the inflation of p–values in the models in the GWAS of the polygenic phenotype, where the credible interval for the 𝜆_gc_ overlapped each other for all heritability rates (**Supplemental Figure 2c**). In the GWAS of hybrid phenotypes, where the environmental noise has non-random structure, 𝜆_gc_ was significantly higher in the PC-adjusted model (𝜆_gc_= 4.498 [CI: 4.355-4.648]) compared to the SPC-adjusted model (𝜆_gc_=1.581 [CI: 1.524-1.635]) for the hybrid smooth phenotypes (**Supplemental Figure 3c**). 𝜆_gc_ for both models increased compared to the scenario with same narrow-sense heritability (h=0.3) and random environmental noise. While SPCs had a lower 𝜆_gc_ in the GWAS of hybrid sharp phenotype (𝜆_gc_=1.321 [CI: 1.287-1.355]) compared to PCs (𝜆_gc_ =1.365 [CI: 1.334-1.396]), their credible intervals overlapped.

To better understand the impact of population structure in the analysis of the polygenic phenotype, we stratified variants into causal and non-causal, referring to variants neither truly associated with the phenotype nor in Linkage Disequilibrium (LD) with causal variants. We assumed that any systematic differences in calculated p-values or effect sizes of non-causal variants in between the two models are likely due to population structure rather than true genetic association. Among the non-causal variants, p-values calculated using PCs were consistently lower (Wilcoxon P-value: 1.54 × 10^−53^), which suggests they are more affected by confounding. However, the mean effect size estimates for these variants were higher for the SPC models; suggesting the lower p-values in the PC model were not due to higher power but may rather reflect differences in how population structure is accounted for. For causal variants and variants in linkage disequilibrium with causal variants, there were no differences in estimated effect sizes using SPCs and PCs.

We used the hybrid phenotypes to evaluate the calibration of each model’s effect size estimate against the ground truth simulated effects. Looking exclusively at estimated effect sizes for variants in high LD (r^2^ > 0.1) with the causal variants in the GWAS of medium heritability phenotype, both models showed similar accuracy in estimating the true simulated effect sizes. (**Supplemental Figure 4a**). Using the same effect sizes along with non-random environmental noise, in the form of the hybrid smooth phenotype, resulted in drastically lower accuracy in the PC-adjusted model, while not significantly affecting the accuracy SPC adjusted model (**Supplemental Figure 4b**). Conversely, when looking at variants in low LD (r^2^ < 0.1) with the causal variants, under random environmental noise, PC-adjusted model found two false-positive variants with a p-value lower than the multiple-testing threshold (p-value < 5 × 10^!$^), whereas the SPC-adjusted model found one. After adding structured noise in the hybrid smooth models, SPC-adjusted model highlighted the same variant, while the PC-adjusted model highlighted 1874 false-positive variants that passed the multiple testing threshold.

To explore the efficacy of SPCs in adjusting for confounding in rare variant analysis, we conducted GWAS of rare variants with all simulated phenotypes. The GWAS of rare variants showed similar 𝜆_gc_ trends to the GWAS of common variants (**Figure 3d**) across all phenotypes, with performance gap decreasing due to lower power, with the exceptions of smooth and hybrid smooth phenotypes. In GWAS of the smooth phenotype, 𝜆_gc_ of the PC model increased significantly to 7.576 [CI: 7.514-7.639], while 𝜆_gc_ of the SPC model exhibited a modest increase to 1.119 [CI: 1.114-1.124]. Similarly, in the analysis of hybrid smooth phenotype, we estimated 𝜆_gc_ values of 5.480 [CI: 5.336-5.630] and 1.162 [CI: 1.157-1.167] for the PC-adjusted and the SPC adjusted models respectively.

As an additional analysis, we used the environmental phenotypes with no underlying causal genetic variation to calculate false-positive rates in the GWAS of common and rare variants. In the GWAS of the smooth phenotype, adjustment using PCs resulted in 1918.9 [std:22.38] false-positives variants passing the Bonferroni multiple testing threshold (5×10^-8^) in the analysis of common variants and 12042.35 [std:71.32] false-positives in the analysis of rare variants. Adjustment by SPCs resulted in 3.6 [std:4.86] and 0.3 [std:0.56] false positives in the analysis of common and rare variants, respectively, suggesting that the SPC method has significantly lower Type I error rates in the presence of population structure confounding. These results demonstrate the robustness of SPC-adjustment to both rare and common allele frequency variants.

### Comparison against alternative strategies of covariate extraction

We next compared SPCs against alternative strategies to calculate fixed-effect covariates to adjust for recent population structure. Previous work by Zaidi and Mathieson demonstrated that PCs derived from rare variants can partially alleviate the limitations of common PCs (Zaidi and Mathieson 2020). In our simulations, SPCs outperformed PCs of rare variants both in terms of predictive performance (R^2^ score) and decreasing the inflation of p-values. While PCs of rare variants had a higher R^2^ score with the environmental phenotypes compared to PCs of common variants, their performance was consistently lower than that of SPCs (**Supplemental Figure 1, Supplemental Table 1**).

Additionally, principal components derived from the IBD relatedness matrix were previously used to improve representation of recent population structure compared to PCs in simulations (Zaidi and Mathieson 2020). In line with those findings, we explored alternative strategies for the calculation of continuous covariates using the IBD relatedness matrix for comparison with SPCs. We calculated 11 additional sets of covariates from IBD relatedness data, comparing principal components to spectral components and exploring a range of possible minimum IBD sharing thresholds in the generation of the relatedness matrix (6cM, 10cM, and 15cM). We found SPCs, calculated using a threshold of 6cM, to be among the best performers in terms of PVE of simulated phenotype (**Supplemental Figure 1**). We also repeated the SPC-adjusted GWAS using two higher IBD sharing thresholds (10cM and 15cM) and found that 6cM threshold yielded 𝜆_gc_ values that were equal or lower than those of higher thresholds (**Supplemental Figure 5**). Overall, our results demonstrate that SPCs provide a more accurate adjustment for recent population structure than alternative covariate strategies (further details in Supplemental Material).

### Evaluating performance in narrow-sense heritability estimation

Finally, we evaluated the impact of SPCs on estimating narrow-sense heritability, the proportion of phenotypic variance that is due to a genetic component (Yang et al. 2017), where adjustment of phenotype values using PCs is a common measure to lower the inflation of estimates due to population structure (Abdellaoui et al. 2019; Cook et al. 2020). We calculated the narrow-sense heritability of the smooth phenotype using the GREML method implemented in the GCTA software package along with a GRM calculated based on common variants. This resulted in a heritability estimate of 1.00, indicating the phenotype is fully heritable genetically (**Supplemental Figure 6**). Adjustment using PCs of rare variants resulted in a heritability estimate of 0.45. However, adjustment using SPCs resulted in a 12-fold decrease of the estimate to 0.08, suggesting a significantly lower genetic heritability component, which is in line with the non-heritability of the phenotype. This illustrates the optimal effectiveness of SPCs against PCs when analyzing phenotypes with strong environmental components.

To test for overcorrection of genetic factors that leads to the underestimation of narrow-sense heritability (Evans et al. 2018), we calculated the heritability of the polygenic phenotypes. Across all heritability values, the estimated credible intervals of narrow-sense heritability for both approaches overlapped with each other (**Supplemental Table 2**). Both approaches yielded heritability estimates similar to the unadjusted heritability estimate, demonstrating that the mixed effects model deployed by GCTA adequately addresses population structure in these scenarios. We simulated another heritable phenotype with high heritability (0.8) and lower polygenicity to explore the effects of lower polygenicity on heritability estimates. There, uncorrected estimated heritability for this phenotype was 0.43 (std:0.04). The estimated heritability after correcting for SPCs did not significantly change (0.42-std:0.04). However, correcting for PCs decreased the heritability estimate to 0.36 (std:0.04) (**Supplemental Figure 7**). Together, this suggests that SPCs have advantages over PCs in accurately estimating heritability in the context of population stratification.

### Measuring the effects of upstream noise

Artifacts of noise and errors in IBD estimation pipelines, including phasing switch errors, false-positive IBD segments, missing IBD segments, and incorrect IBD segment boundaries negatively affect the quality of IBD estimation and, consequently, global sharing estimates. For example, we previously showed that the boundaries of IBD segments might be underestimated (Shemirani et al. 2021). We measured the effects of these artifacts on the performance of SPCs in terms of PVE by comparing SPCs derived from inferred IBD (estimated SPCs) against SPCs calculated using ground-truth IBD data extracted directly from the simulated ancestral recombination graphs (ARGs; c.f. Methods). We calculated the PVE in the hybrid smooth phenotype by the first 25 PCs, SPCs, and ground-truth SPCs (**Supplemental Figure 8**). While ground-truth SPCs outperformed the estimated SPCs, their performance was qualitatively similar, with PVEs of 0.59 vs. 0.58 for ground-truth and estimated SPCs, respectively, suggesting that upstream artifacts do not cause serious hindrance to the performance of SPCs.

### Analysis of the UK Biobank

We then sought to evaluate the performance of SPCs in real data using genetic data from individuals with self-reported White British ancestry in the UK Biobanks (UKBB; N∼420,000). We focused on three continuous phenotypes (**Figure 3a**): Easting coordinates of birthplace (eastings), which display a linear geographical cline and have negligible true heritability, Body Fat Percentage (BFP), which is established to have medium heritability (h^2^∼0.3), and height, which is established to have high heritability (h^2^∼0.7; Karczewski et al. 2024). In terms of proportion of variance explained, we saw similar results as the simulated data, with SPCs outperforming PCs for all phenotypes. The difference was particularly significant for eastings. The GREML narrow-sense heritability estimates revealed similar patterns, though the gap in estimates was smaller compared to the simulated analysis. The heritability analysis for height and BFP resulted in the same estimates in both models (0.7±0.01 for height, ∼0.31±0.01 for BFP), while the estimates for eastings were 0.63 (std: 0.01) and 0.04 (std:0.01), using PCs and SPCs, respectively. This distinction between easting and the other two phenotypes is similar to that of environmental smooth and polygenic phenotypes in the simulations, respectively.

Next, we conducted GWAS for all 3 phenotypes to measure the inflation of the test statistics due to population structure, using the first 25 PCs and SPCs, age, age squared, sex, and an age/sex interaction variable, as covariates. To account for indirect genetic effects, and cryptic relatedness, which were not present in the simulated phenotypes, we used the BOLT-LMM software, which applies a Bayesian Shrinkage Prior to adjust for those effects (Loh et al. 2015). In the GWAS of eastings, SPCs resulted in a lower inflation of p-values, as evidenced by a lower genomic inflation factor compared to PCs: 1.200 and 1.369, respectively. In contrast, the GWAS of height and BFP showed no significant differences in the inflation of p-values as both approaches yielded the same inflation factor (**Figure 3c**). We then conducted a GWAS of rare variants using imputed genotype data, using BOLT-LMM. Looking exclusively at the imputed rare variants (MAF < 0.01), we observed lower genomic inflation factor values for both models in all three phenotypes along with similar patterns as the non-imputed GWAS, with SPCs having a lower inflation factor (1.147) compared to PCs (1.311) in easting and both models having the same inflation factor for height and BFP.

We then sought to explore the differences between the effect sizes estimated by each model. LD score regression found identical heritability estimates from both models in all phenotypes (**Figure 4**), with a higher intercept for PCs in eastings GWAS (1.362 vs. 1.172). This suggests the difference in performance in eastings was mostly driven by population structure confounding in the PC model (Bulik-Sullivan et al. 2015). For height and BFP, aggregate analysis revealed negligible differences, with SPC-adjusted slope being lower but not significantly. We explored this further for height by extracting the list of loci for which the estimated single variant effect sizes differed significantly between the two approaches. We found that the minor alleles in these loci tended to be more uncommon, compared to all tested variants (Kolmogorov-Smirnov test p-value:5 × 10^−83^), highlighting that the impact of SPCs is most pronounced at lower allele frequencies. The same pattern was also observed in the results for the eastings and BFP GWAS.

**Figure 4.**
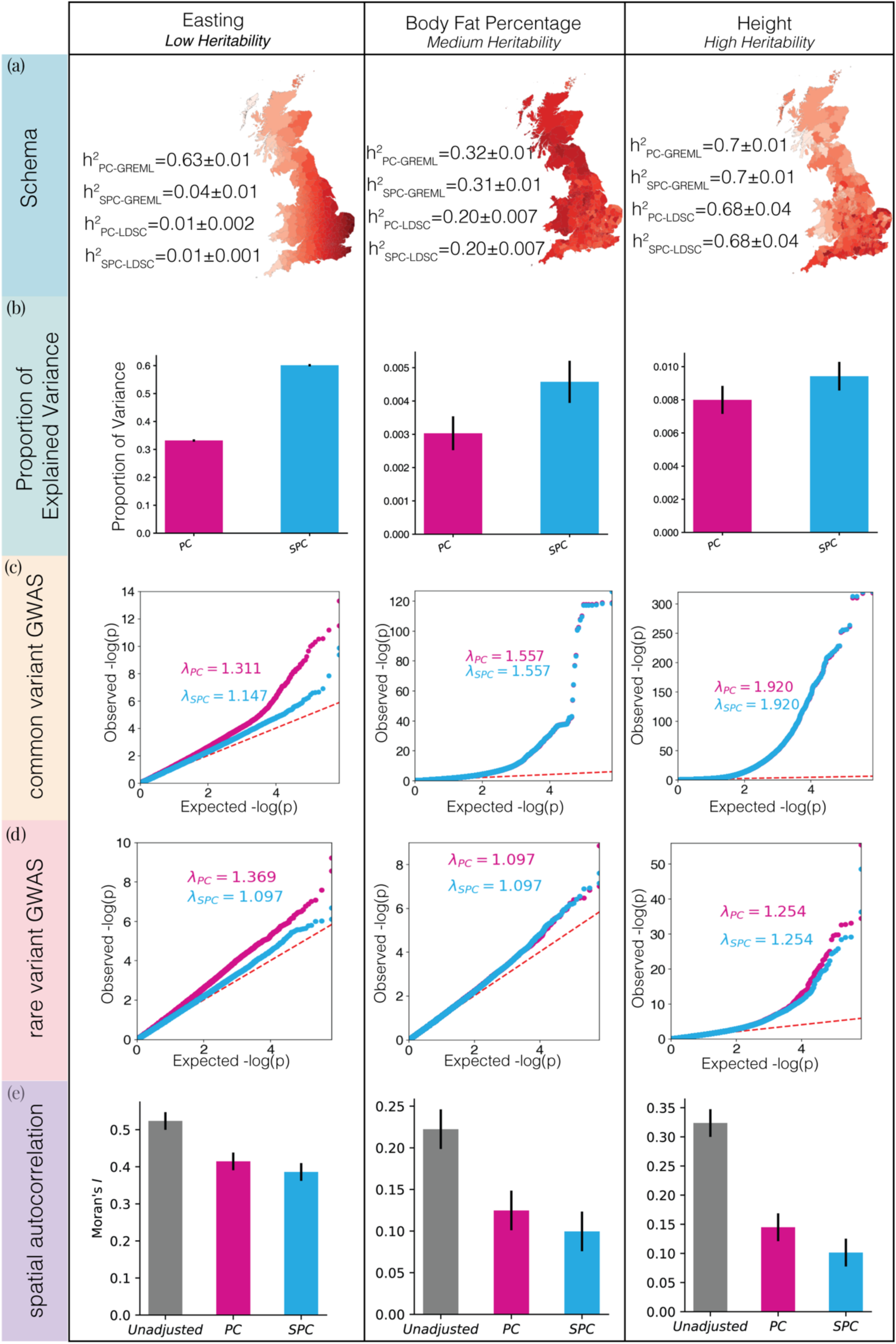
Comparison of PCs and SPCs as covariates to correct for population structure on the outcomes in the UK Biobank. **a)** The structure of each phenotype. We analyzed 3 phenotypes with varying degrees of true heritability, increasing from left to right. Heritability estimation using GREML and LDSC methods, after correcting for recent population structure using both approaches is shown, along with display of average phenotype values in each administrative county in the UK. **b)** Proportion of explained variance in each phenotype by a model that only includes PCs or SPCs as covariates. Error bars represent standard deviation. **c)** QQ plot of the distribution of p-values in the GWAS of common variants for each phenotype that measure the ability of covariates to deal with the effects of recent population structure on the outcomes. **d)** QQ plot of the distribution of p-values in the GWAS of rare variants (MAF < 1%) in the imputed dataset. **e)** Spatial autocorrelation of each phenotype measured in terms of Moran’s I as a comparison of the ability of each set of covariates to correct against the artifacts of environmental effects.

We calculated spatial autocorrelation of the phenotypes, in the form of Moran’s I, as a measure of indirect environmental effects (Moran 1948). A previous study showed the spatial autocorrelation of phenotype distributions is reduced after adjustment using PCs, illustrating the efficacy of PC adjustment in addressing confounding due to such effects (Abdellaoui et al. 2019). Here we replicated that experiment and added SPC adjustment (**Figure 4e**) as another comparison approach. The Moran’s I for all three phenotypes decreased after adjustment using PCs, 0.52 to 0.41 for easting, 0.22 to 0.12 for BFP, and 0.32 to 0.15 for height. Adjustment using SPCs further reduced Moran’s I estimates to 0.38 for easting, and 0.10 for BFP and height. In summary, our analysis demonstrates that SPCs consistently outperform PCs in reducing population structure confounding, particularly for phenotypes with strong environmental components, and show comparable performance for highly and moderately heritable traits like height and BFP, especially when considering rare variant effect sizes and spatial autocorrelation.

As a sensitivity analysis, we conducted a comparison of SPCs against other covariate extraction techniques by evaluating their PVE on a random subset of the UK Biobank participants (N=50,000; **Supplemental Figure 9**). SPCs outperformed both PCs of rare and common variants. We note that in this dataset (unlike in our simulations), PCs of rare variants could not outperform PCs of common variants. We then calculated additional component sets based on minimum IBD sharing thresholds (6cM, 10cM, and 15cM), graph weighting schemes (weighted vs. unweighted graphs), and component calculation methods (spectral vs. principal components of IBD sharing), resulting in 12 sets of covariates (including SPCs), replicating our analysis using simulated data. For each sharing threshold, SPCs outperformed their alternatives, highlighting the ability of spectral components in extracting localized, fine-scale signals of population structure, along with the advantages of utilizing unweighted matrices, especially in the first 5, and the first 10 dimensions. We further showed how a higher sharing threshold can negatively impact PVE by excluding a significant proportion of IBD signals (**Figure 5**). Together with our simulated results, these analyses demonstrate that SPCs, calculated at the 6cM threshold, provide a more accurate adjustment for recent population structure than alternative adjustment strategies (further details in Supplemental Material).

**Figure 5.**
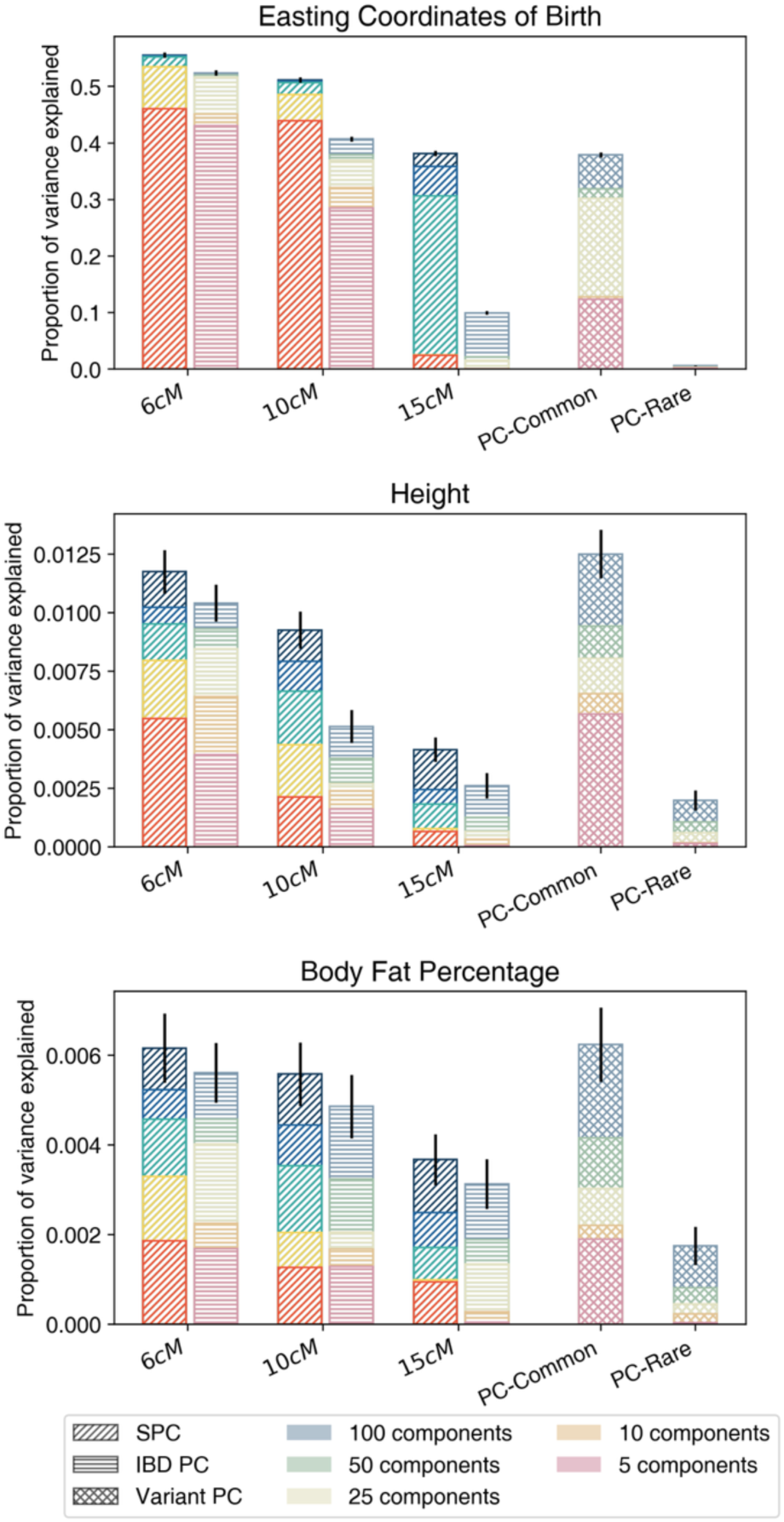
Bar plots of the PVE in eastings, height, and BFP explained by SPCs (derived from binary IBD relatedness), IBD PCs (derived from binary IBD relatedness), and PCs (derived from genetic variants) in the UK Biobank, Exploring the effects of minimum lBD sharing parameter, and decomposition approach on the performance of covariates. Principal components of common and rare variants are displayed on the rightmost side of the figure. For other components, labels on the x-axis indicate the minimum IBD sharing threshold used to generate the components. Spectral (SPC) and principal (IBD PC) components were calculated per relatedness graph, indicated by diagonal and horizontal shading lines, respectively. SPCs, are highlighted in brighter colors. PCs of common and rare variants are subscripted as PC-Common and PC-Rare, respectively.

To measure the robustness of SPCs in the presence of ascertainment bias, we explored the effects of network sparsity on the SPCs. In PC calculation, a lack of balance in representation may result in the PC’s inability to capture population structure signals from subpopulations with lower representation (A. B. Lee et al. 2010). The IBD analysis of the UKBB cohort, due to its demographic history, leads to a densely connected graph, where most individuals have hundreds of IBD connections to others (**Supplemental Figure 10**). However, imbalances in representation result in a limited number of participants with lower connectivity. Here, connectivity (or node degree) is the number of edges per node. We searched for individuals with the lowest connectivity levels in a random subset (N=50,000). The lowest 20 node degrees ranged from 4 to 131 (mean = 84.65; median = 103). We looked at the distribution of SPC components to see if the nodes with low connectivity cluster near zero due to lower variability in their latent representation, suggesting failure to capture their underlying structure. We calculated the variance of each SPC dimension and compared them to the variances of a random sample of the same size (**Supplemental Figure 11**) and found that the variances calculated for lower connectivity nodes were not significantly lower than those of the random sample. When applying this to principal components, low-connectivity nodes exhibited significantly reduced variance. The distributions across SPC dimensions further illustrate SPCs’ robustness to ascertainment bias (**Supplemental Figure 12**).

## Discussion

We introduced SPectral Components (SPCs), a graph-based approach that leverages IBD graphs to capture localized, non-linear fine-scale population structure, and transforms them into a continuous representation for statistical genetics analyses. This method overcomes some limitations of Principal Components (PCs), which are optimized for modeling continuous linear ancestral relationships, but struggle to fully account for recent demographic events that shape patterns of rare variation. Our results demonstrate that SPCs outperform PCs, explaining over 90% of fine-scale population structure in simulations while reducing genomic inflation in genome-wide association studies (GWAS) for both environmental and heritable phenotypes. Notably, SPCs improved rare variant association analyses by reducing genomic inflation, while providing more accurate heritability estimates and avoiding overcorrection for the highly heritable trait of height. Spatial autocorrelation analysis further confirmed that SPCs better control confounding from environmental effects, reducing Moran’s I more effectively than PCs. These findings establish SPCs as a powerful and scalable tool to improve population structure adjustment and confounding control in large-scale, diverse biobank studies.

This work builds on previous research on IBD- or rare variant-based GRMs which have shown that the latter models can often overcorrect for population structure, leading to loss of true signal in genetic analysis. For example, several studies (Browning and Browning 2015; Evans et al. 2018) demonstrate that overcorrection can occur because IBD GRMs disproportionately emphasize recent, localized relatedness, which can obscure genuine associations. Additionally, it extends prior applications of spectral approaches to common variant data, which demonstrated they can be more robust than PCs in finding hidden population structure (A. B. Lee et al. 2010). However, those applications use variants that are shared identical-by-state, rather than haplotypes shared identical-by-descent, which lack genealogical context and are expected to have poorer resolution of confounding for recent population structure and rare variant association. The distinction between fixed effects and random effects on these models further complicates the adjustment process. Fixed effects, like PCs, explicitly control for population structure by removing specific sources of variation, whereas random effects models, like those incorporating GLMMs, aim to partition variance across all genetic relationships, but may inadvertently attenuate rare variant signals by treating them as background noise. SPCs provide a middle ground by capturing fine-scale population structure without overcorrection, using a fixed-effect approach tailored to discrete, localized signals, while maintaining the broader interpretability of linear mixed models. Moreover, the use of SPCs showed a clear reduction in genomic inflation for rare variants, therefore providing a significantly more robust adjustment. Other work on alternative approaches, such as using ARGs to infer genome-wide genealogies that can be combined with GLLMs to improve power for rare variant association has recently been shown to scale to large biobank datasets (Nait Saada et al. 2020; Zhang et al. 2023; Gunnarsson et al. 2024; Christ et al. 2024). While these approaches show promise, they currently have some limitations, such as implicating large genomic regions in variant association and requiring methodological improvements to LMM scalability. Future work could explore the use of PCs, ARGs, and SPCs in tandem as a powerful tool for addressing the challenges posed by population structure in modern genomics (Zaidi and Mathieson 2020; Ma and Shi 2020).

Another focus of this study was to examine how SPCs capture environmental effects, which often overlap with recent population structure (Zaidi and Mathieson 2020). The increased accuracy of SPCs compared to PCs is especially important given the increasing recognition of environmental confounding in GWAS, where geography and patterns of recent demography can lead to spurious associations. In this context, SPCs explain a significantly greater proportion of variance for environmental phenotypes, such as easting coordinates. This result aligns with earlier work (Abdellaoui et al. 2019; Cook et al. 2020; Leslie et al. 2015; Zaidi and Mathieson 2020), highlighting the difficulty of decoupling environmental and genetic factors in geographically clustered populations. Our findings support this previous work, demonstrating that population structure confounding remains a major challenge even in large-scale homogeneous cohorts like the White Britons from the UK Biobank. We showed this for the highly polygenic trait of height, as demonstrated by measuring the spatial autocorrelation in the form of Moran’s I. SPCs were able to further decrease the spatial autocorrelation of height compared to PCs. Our results also support the use of SPCs in heritability analysis, where adjustment using SPCs reduced genetic heritability estimates by accounting for environmental confounding without overcorrection. By improving adjustment for the environmental confounding effects, SPCs provide a critical advance in the accuracy of genomic analysis for phenotypes that are heavily influenced by recent demography or environmental variables.

We highlight several limitations to this study and directions for future development. First, our study was restricted to unrelated samples of White British descent in the UK Biobank. SPCs should be further evaluated in more diverse datasets where relatedness, cryptic family structure, admixture, and population differentiation may introduce additional challenges. Second, though SPCs consistently outperform PCs in this study, additional work is needed to further validate the power of SPCs across settings such as smaller cohorts with sparser graphs, phenotypes with lower heritability, and dichotomous outcomes where collinearity may be more pronounced. Third, further work could extend SPCs to additional analytic frameworks, like burden testing, generalized additive models, and polygenic risk score calculation, to assess their ability to improve prediction accuracy and calibration. Fourth, while our analysis focused on SPCs from global IBD graphs, alternative strategies merit exploration, including clustering-based partitioning of IBD graphs and multi-segment IBD detection approaches (Temple and Thompson 2025) which may capture additional fine-scale relatedness. Finally, SPCs could also be evaluated as covariates in IBD-based mapping of quantitative trait loci, and their ability to resolve fine-scale population structure differences suggest they may help improve mapping accuracy (Cai and Browning 2025; Chen et al. 2023).

In summary, SPCs offer a robust, continuous alternative to PCs for adjusting for recent population structure in genomic studies. By capturing fine-scale, non-linear demographic patterns, SPCs provide more accurate adjustment for confounding due to environmental effects and recent ancestry, particularly in large-scale cohorts. As large-scale biobanks continue to grow in size and diversity, the impact of rare variants in genetic analysis will become increasingly pronounced. In this context, tools like SPCs will be essential for ensuring accurate results, particularly as researchers delve deeper into the genetic architecture of rare variants on common disease and traits influenced by population-structured dynamics. By addressing the combined challenges of population structure, environmental confounding, and rare variant associations, SPC-based approaches pave the way to uncover novel insights into complex disease architecture.

## Methods

### Calculating Spectral and Principal Components

The calculation of SPCs can be described in 4 main steps (**Figure 1**).

*Step 1:* We conducted QC and phasing on array data. The phased data was used by the IBD estimation algorithm, iLASH, to detect pairwise for IBD for each dataset. An in-depth explanation of genotype QC and phasing, along with details about running iLASH and estimating IBD is described in (Shemirani et al. 2021). Throughout this paper we use the term IBD to refer to long haplotype segments (≥6cM) that are shared identical-by-state (IBS) and, given their length, are extremely likely to be inherited from a recent common ancestor. This operational definition is consistent with prior work (Browning and Browning 2015; Shemirani et al. 2021; Gusev et al. 2009). We acknowledge that some definitions of IBD differ, and we have added this clarification to reduce ambiguity. The results of the first step can be represented as 𝑆 the set of all IBD segments, long enough to be detected. Each segment has a starting point and an end point 𝑠_$_, 𝑠_%_, located on autosomal chromosome k.

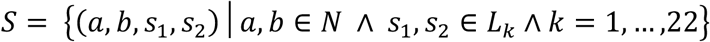

*Step 2:* the aggregated sums of pairwise IBD sharing (i.e. the total or combined length of IBD haplotypes that are shared per pair of individuals) were calculated and used to generate an IBD relatedness graph. On this graph, nodes represented participants and edge weights represented the aggregated IBD sharing. A minimum threshold of aggregated IBD sharing is enforced both as a QC measure (Belbin et al. 2021) and to reflect the population structure of the dataset.

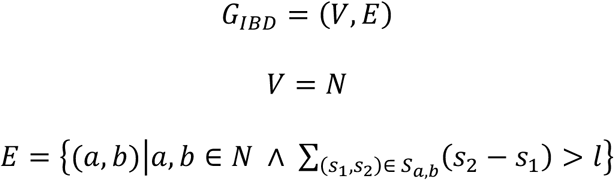

To calculate SPCs we used a threshold of 6cM. However, we explored two other thresholds of 10cM and 15cM for comparison (c.f. Supplementary Material).

*Step 3:* The 𝑛 × 𝑛 adjacency matrix 𝑨 was generated, representing the relatedness graph, where rows and columns both represent the 𝑛 participants in the dataset. Each element 𝑎_12_ in this matrix is equal to 1 if the aggregated length of IBD segments shared between the two participants 𝑖 and 𝑗 on the graph is above the threshold of 𝑙 = 6𝑐𝑀; otherwise, the value of the elements is set to zero.

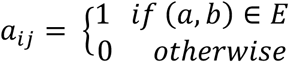

We refer to this matrix as the binary IBD relatedness matrix. We also explored the use of non-binary relatedness matrices for comparison due to their similarity with common Genetic Relatedness Matrices (GRM; c.f. Supplementary Material).

*Step 4:* Any graph adjacency matrix can be transformed into a Laplacian form (Shuman et al. 2013). Here we defined a symmetrically normalized Laplacian:

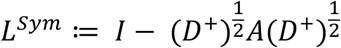

Where 𝐼 is the identity matrix, 𝐷 is defined as the degree matrix, a diagonal matrix where each element 𝑑_11_ is equal to the total number of neighbors for participant 𝑖, and 𝐷^5^is the (Moore-Penrose) inverse of 𝐷. The Eigenvectors of the Laplacian matrix are commonly referred to as the spectral components of the graph. Briefly, SPCs are the spectral components of the binary IBD relatedness graph with a cut-off threshold of 6cM. Thus, akin to PCs, SPCs attribute a set of numerical values to each vertex based on its projections on the set of principal axes of variation in the graph. The level of detail represented by each axis depends on the corresponding eigenvalue association with it. Eigenvector associated with the smaller eigenvalues will assign similar number to neighbors on the graph. As the associated eigenvalue increases, similar values are assigned to participants that are incrementally further from each other on the graph (further details in Supplemental Material).

We have implemented this pipeline as a Python library that transforms flat text IBD graph files to SciPy sparse matrices and uses CPU or GPU acceleration to calculate SPCs using the same library.

### Generating simulated data

We used the MSprime python library to implement the simulation pipeline to ensure the distribution of IBD connections and their length is concordant with real datasets (Baumdicker et al. 2022). Following guidelines on simulation of homogeneous population with recent structure akin to that of UK Biobanks (Zaidi and Mathieson 2020), our pipeline simulated N-by-N grids of demes with constant migration rate of 0.05. We set N=5 (25 demes) for our main simulation of chromosome 1 and 50,000 diploid individuals. We also simulated a replication dataset with N=4 with 22 chromosomes and 8,000 diploid individuals. The length of the chromosomes in the 8,000-sample simulation were adopted from the HapMap project (International HapMap Consortium et al. 2007). Chromosome specific recombination rates were adopted from the stdpopsim library metadata (Adrion et al. 2020). In the 50,000-sample simulation, we used the same library to obtain chromosome 1 specifications, with 248,956,422 base pairs and recombination rate of 1.149 × 10^−8^. The migration rate of 0.05 has been shown to establish demes with average divergence, measured by *F_st_*, equal to that of neighboring administrative counties in the UK Biobank dataset (Leslie et al. 2015). All demes originate from a single ancestral population 300 generations ago. The first 150 generations going back from the current day were simulated using Discrete Time Wright-Fisher (DTWF) model, implemented in the MSprime library to ensure IBD length distribution is similar to that observed in human genotype datasets. The simulation pipeline is available publicly on GitHub.

While calculating true IBD data is readily possible with MSprime, we sought to replicate common noise present in real-world applications, such as switch errors and false-positive IBD segments. The simulated tree sequences were converted to whole-genome sequence data in PLINK 1.9 format in order to delete the ground-truth phase information. The data was then filtered to retain only common variants (MAF>1%) and then phased using Eagle v4.2 with default parameters. Genetic map files for Eagle were generated using the recombination rates applied in the simulations. We have provided the code to generate these files in our simulation GitHub repository. IBD segments were estimated using the iLASH software v1.0.1 with default parameters (min_length 2.9, perm_count: 20, shingle_size: 15, shingle_overlap: 0, auto_slice:

True, cm_overlap: 1, bucket_count: 5, match_threshold: 0.99, interest_threshold: 0.70). For the comparative analysis of downstream applications, 7 types of phenotypes were simulated (Figure 3a). The first category, referred to as environmental smooth phenotypes, are non-heritable phenotypes with a distribution that depends on the deme of origin. All values were generated using a normal distribution with constant variance across all demes. Moving vertically from top to bottom, the mean of the normal distribution decreases monotonically per row. It decreases from a value of 2𝝈, for the top row, to zero, for the bottom row. Briefly, in a 5 by 5 grid simulation, this means the phenotype values on the top row are drawn from the distribution 𝑦∼ 𝒩(2𝜎, 𝜎), the values on the second row are drawn from the distribution 𝑦∼ 𝒩(1.5𝜎, 𝜎), each subsequent row has a mean that is 0.5𝜎 smaller than the previous, with last row having a mean of zero. Second phenotype, called the environmental sharp phenotype is also non-heritable with a normal distribution with a non-zero mean (𝑦∼ 𝒩(𝜇, 𝜎)) at the target deme and a zero-centered normal distribution (𝑦∼ 𝒩(0, 𝜎)) in all other demes. We used a 𝜎 = 1 in our analysis for this manuscript. We generated 3 heritable polygenic phenotypes, to which we refer, based on their heritability, polygenic – low heritability (h^2^=0.1), polygenic – medium heritability (h^2^=0.3), polygenic high heritability (h^2^=0.08). For these phenotypes, we selected causal SNPs randomly from each window of 10 KB along the genome (24,813 causal variants per phenotype) with effect sizes drawn from 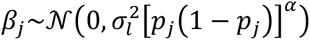 where 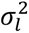 is calculated based on selective pressure parameter 𝛂=-0.5, and the total heritability of the phenotype (𝜎_+_ = 0.1, 0.3, and 0.8) via the formula 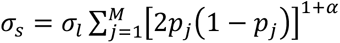. We then used the weights along with the simulated whole genome sequence data, and the score calculation functionality in the PLINK software (--score) to generate the mean phenotype values for each individual. The remaining variance of the phenotypes were added using a random normal distribution with variances of 0.9, 0.7, and 0.2 respectively.

Finally, we simulated two hybrid phenotypes by combining the environmental smooth and sharp phenotypes and the polygenic phenotype with medium heritability (h^2^ = 0.3), by reweighting the environmental phenotype to have a variance of 0.7 and adding the values of the two phenotypes to generate hybrid smooth and hybrid sharp phenotypes, respectively.

LD data to detect variants tagging causal ones was generated using PLINK with parameters: “--list-all, --show-tags, --tag-kb 2000, --tag-r2 0.1.

Ground-truth IBD data were extracted using the ibd_segment function in the tskit library with a minimum span of 3cM and maximum TMRCA of 200.

### UK Biobank Data

Phased genotype data along with the imputed data for the UK Biobank project was downloaded through the accession code 84541. The UK Biobank participants were genotyped using the Applied Biosystems UK BiLEVE Axiom Array by Affymetrix 807,411. The genotype data was phased using the SHAPEIT3 software and the 1000 Genomes Project as the reference panel (Bycroft et al. 2018; Auton et al. 2015; Delaneau et al. 2019). After removing indels from the 22 autosomal chromosomes, a total of 655,532 SNPs were retained. Data field 21000, based on self-reported race/ethnicity was used to extract (N=471,931) individuals with White British ancestry. Information on allele frequency and missingness in both phasing and QC process for GWAS data is available in (Loh et al. 2018). We estimated fine-scale IBD sharing using the iLASH software (Shemirani et al. 2021) using the same parameters mentioned in the previous section. Second degree relatives were identified using KING (parameters: --related --degree 2; N=24,799). From each pair of these relatives, at least one individual was excluded from the analysis (N=405,508). Height information was accessed using data field 20 (N=352,254). BFP was accessed using data field 23099 (N=346,655). Easting coordinates of birth was accessed using data field 129 (N= 343,845). PCs were calculated using PLINK2 software.

### Genome-wide association study

The GWAS of simulated data was conducted using the GCTA software with default parameters and FastGWA option to run linear regression (Jiang et al. 2019). For the common variant analysis, we excluded variants with MAF<1% using PLINK2 software. For the rare/uncommon variant analysis we excluded variant with MAF>1% using the same software. The GWAS in UK Biobank was conducted using the BOLT-LMM software version 1 to account for cryptic relatedness and direct genetic effects, absent in the simulated phenotype analysis. Age, Age squared, age, sex, age by sex, and age squared by sex were used as covariates (Loh et al. 2015). The LD reference panel was calculated using genotype data from individuals with reported European ancestry in the 1000 Genomes Project (https://broad-alkesgroup-ukbb-ld.s3.amazonaws.com; access date January 2023). The default genetic map file available in the SHAPEIT software package (Delaneau et al. 2019) calculated for Human Genome version 19 (hg19) was used for genetic distances. Minor allele frequency of 0.001 and a minimum INFO score of 0.3 filters were used for the imputed data.

### Computing Environment

All analyses in this manuscript were performed on the High-Performance Computing Cluster at Mount Sinai. The compute node used featured an Intel Emerald Rapids 8568Y+ CPU with 96 cores at 2.3 GHz (20 cores were used for our applications), and 1.5 TB of RAM (approximately 100 GB were used during our runs). The operating system was CentOS v7.0, with GCC v8.3.0 and Python v3.5.

### Software availability

A python implementation of the SPC algorithm is available at https://github.com/roohy/spc Our simulation pipeline is available at https://github.com/roohy/msprime_ancestry_simulation The Eagle phasing software is available at: https://alkesgroup.broadinstitute.org/Eagle/ The PLINK2 software is available at: https://zzz.bwh.harvard.edu/plink/plink2.shtml The iLASH software is available at: https://github.com/roohy/iLASH

GCTA software is available at: https://yanglab.westlake.edu.cn/software/gcta/

BOLT-LMM software is available at: https://alkesgroup.broadinstitute.org/BOLT-LMM/

## Data availability

Simulated genotype data, along with phenotypes is available on Mendeley repository with doi:10.17632/9dcpdbzv4m.1

## Data Availability

UK Biobank data is available for download through application from https://www.ukbiobank.ac.uk.
LD score reference data was downloaded from https://broad-alkesgroup-ukbb-ld.s3.amazonaws.com.
Simulated genotype data, along with phenotypes is available on a Mendeley repository: 10.17632/9dcpdbzv4m.1
All data produced in the present study are available upon reasonable request to the authors

## Acknowledgements

Conceptualization by R.S., E.E.K., C.R.G., and N.Z. Methodology developed by R.S., and E.E.K., and N.Z., Conceptualization of the simulation of population structure by R.S., and G.M.B., Formal analysis by R.S., E.E.K., and N.Z., reviewing and editing by all authors, and supervision by E.E.K.

This work was supported by the National Human Genome Research Institute of the National Institutes of Health under award numbers R01HG011345 and U01HG010971.

This work was supported in part through the computational and data resources and staff expertise provided by Scientific Computing and Data at the Icahn School of Medicine at Mount Sinai and supported by the Clinical and Translational Science Awards (CTSA) grant UL1TR004419 from the National Center for Advancing Translational Sciences. Research reported in this publication was also supported by the Office of Research Infrastructure of the National Institutes of Health under award number S10OD026880 and S10OD030463. The content is solely the responsibility of the authors and does not necessarily represent the official views of the National Institutes of Health.

