## Supplemental Material for "SPC: a SPectral Component approach leveraging Identity-by-Descent graphs to address recent population structure in genomic analysis"

### Supplementary Information for “SPC: a SPectral Component approach to address recent population structure in genomic analysis”

#### Supplemental Figures

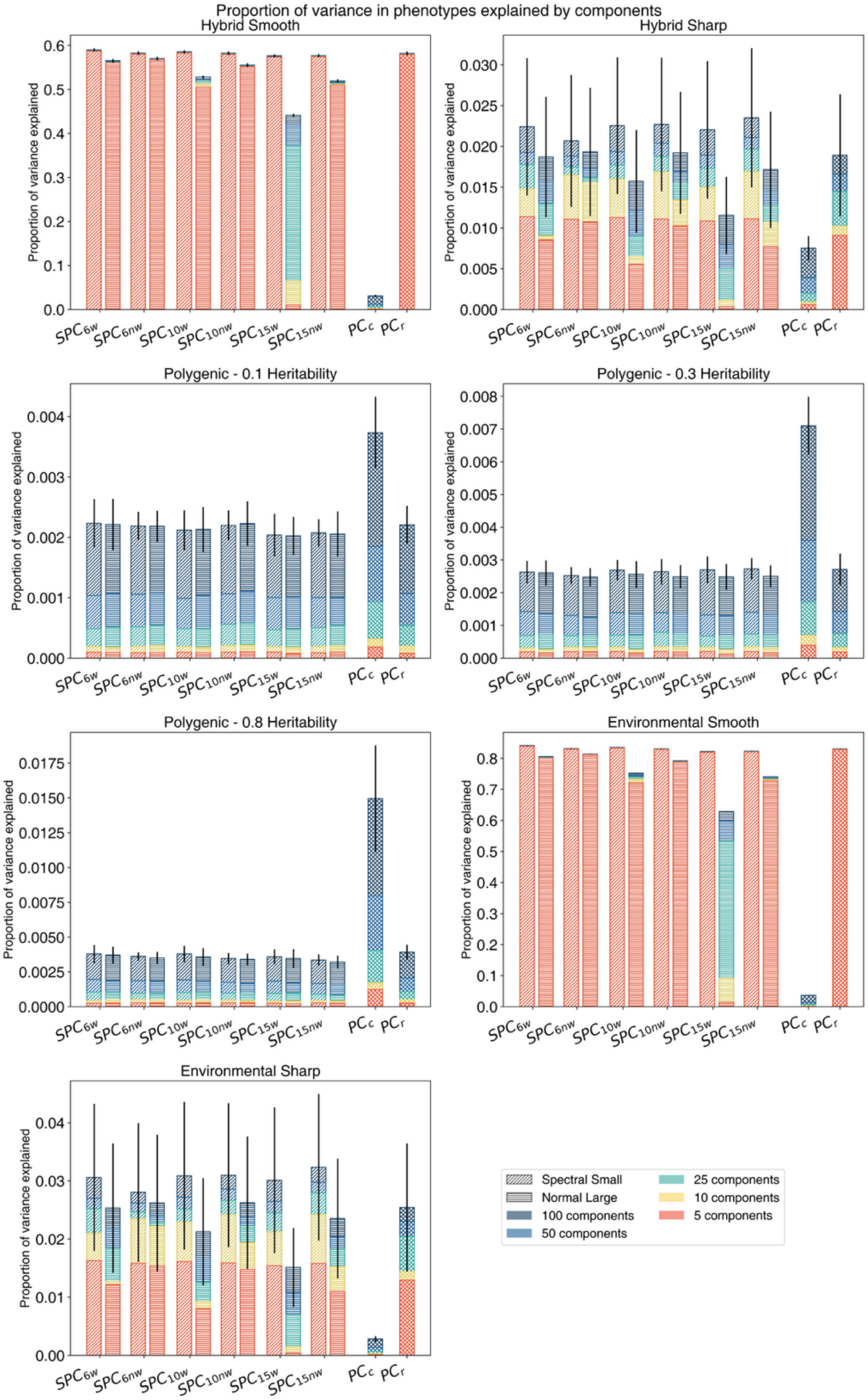


**Supplemental Figure 1 –** Proportion of variation in the phenotypes explained by the principal components in 7 simulated phenotype groups. Principal components of common and rare variants are displayed on the rightmost side of the figure. For other components, labels on the x-axis indicate the data type used to generate the components. The minimum length of IBD used to generate the components (6cM, 10cM, 15cM) are denoted in the subscript. Components calculated from weighted graphs are denoted with 'w' in the subscript, and those calculated from the binary graphs are denoted with ‘nw’. Spectral and principal components of each data type were calculated, indicated by diagonal and horizontal shading lines, respectively.

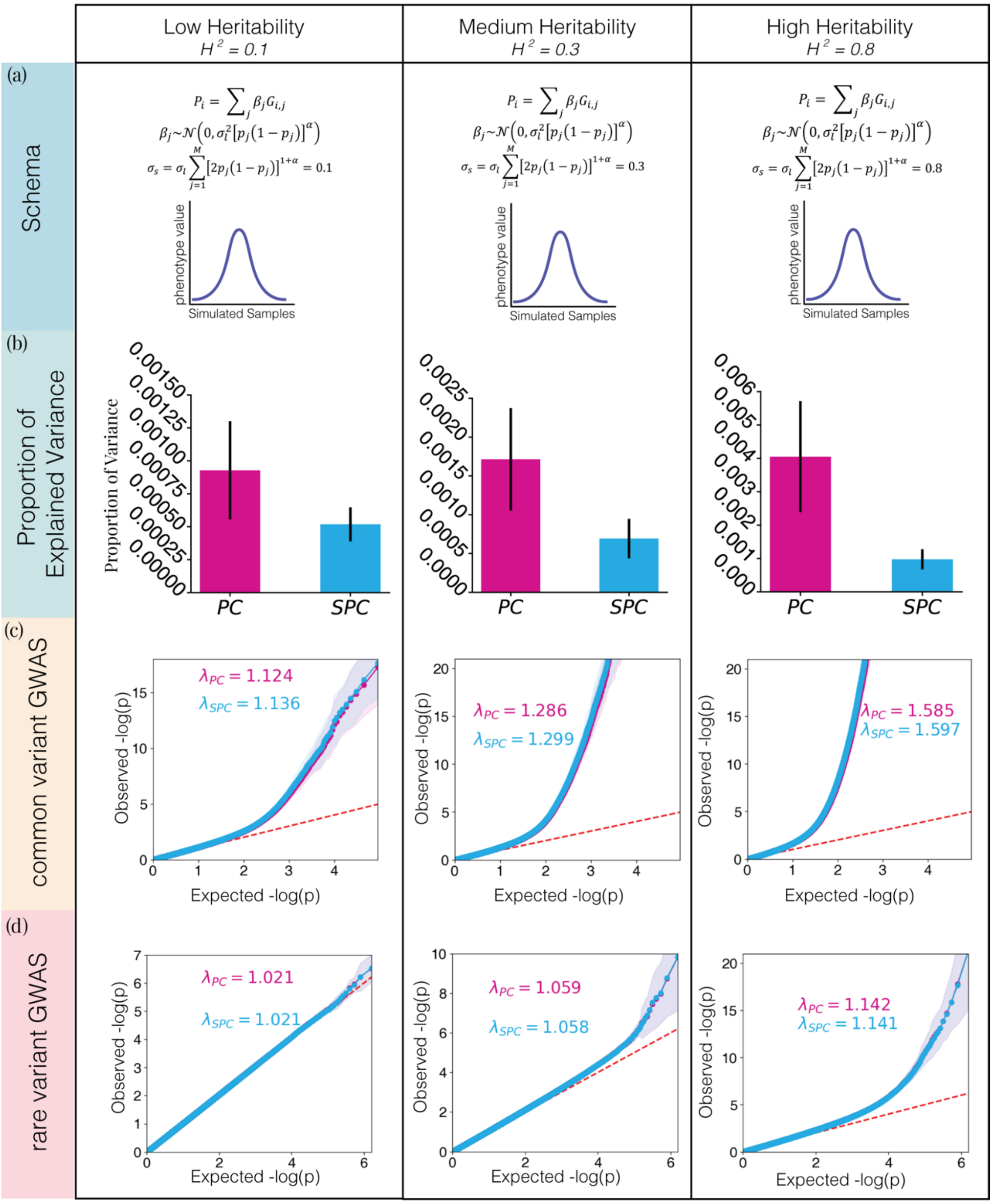


**Supplemental Figure 2 –** Comparison of PCs and SPCs as covariates to correct for population structure on the outcomes in simulations *of polygenic heritable phenotypes.* **a)** The schema of simulated phenotypes. These polygenic phenotypes were determined by assigning effect sizes to causal variants, randomly selected from windows of 10,000 base pairs, and calculating the polygenic score based on the effect sizes. Effect sizes were drawn from a normal distribution with a mean of zero and a variance derived from minor allele frequency, heritability, and selective pressure. $\sigma_s$ determines the expected heritability of the phenotype, while $\alpha$ determines the selective pressure, with negative values resulting in higher effect sizes assigned to variant with low minor allele frequency. Tested values for $\sigma_s$ were: 0.1, 0.3, and 0.8. $\alpha$ was set to -0.5 across all experiments **b)** proportion of variation in the phenotypes explained by the first 25 PCs and SPCs in each phenotype. The error bars represent the bootstrapped standard deviation calculated from 2,000 repetitions. **c)** Genomic inflation of the results of GWAS analysis of simulated phenotypes using common variants only. **d)** Genomic inflation of the results of GWAS analysis of simulated phenotypes using rare variants (10<MAC<500).

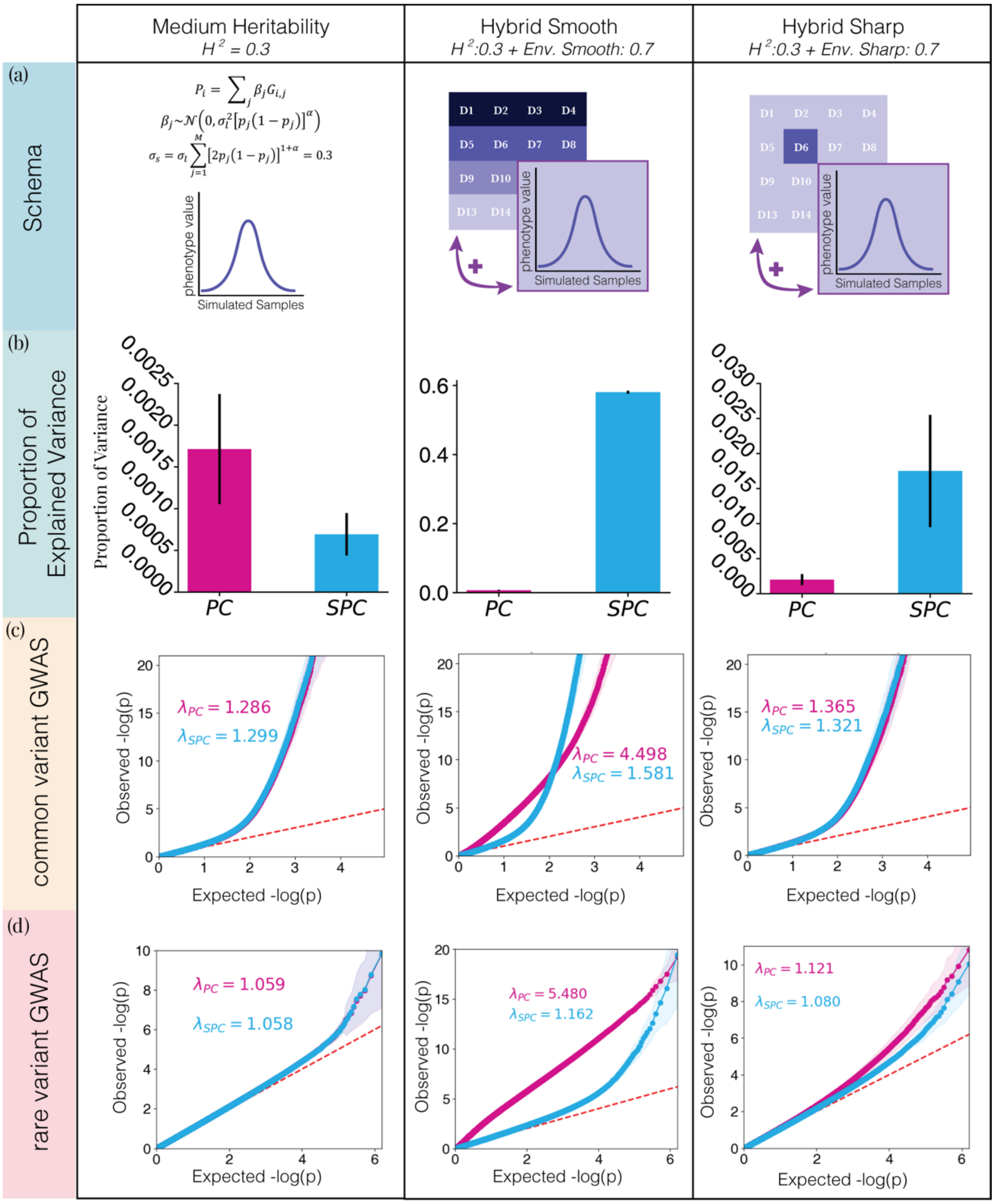


**Supplemental Figure *3* –** Comparison of PCs and SPCs as covariates to correct for population structure effects on phenotypes in simulations *of polygenic heritable phenotypes with structured environmental noise.* **a)** The schema of simulated phenotypes. First, all three phenotypes share the same underlying polygenic basis. This polygenic phenotype was determined by assigning effect sizes to causal variants, randomly selected from windows of 10,000 base pairs, and calculating the polygenic score based on the effect sizes. Effect sizes were drawn from a normal distribution with a mean of zero and a variance derived from minor allele frequency, heritability, and selective pressure. $\sigma_s$=0.3 determines the expected heritability of the phenotype, while $\alpha = -0.5$ determines the selective pressure, with negative values resulting in higher effect sizes assigned to variant with low minor allele frequency. The remaining variance (0.7) in each phenotype is derived from normally distributed random noise functions. In the phenotype with medium heritability, all samples draw from the same zero-mean normal distribution. In the hybrid smooth phenotype, the mean of the distribution depends on the horizontal coordinates of demes. The top row has a mean of 1.4, while the bottom row has a mean of zero. In hybrid sharp phenotype, only one deme draws its environmental effects from a non-zero mean normal distribution. **b)** proportion of variation in the phenotypes explained by the first 25 PCs and SPCs in each phenotype. The error bars represent standard deviation. **c)** Genomic inflation of the results of GWAS analysis of simulated phenotypes using common variants only. **d)** Genomic inflation of the results of GWAS analysis of simulated phenotypes using rare variants (10<MAC<500).

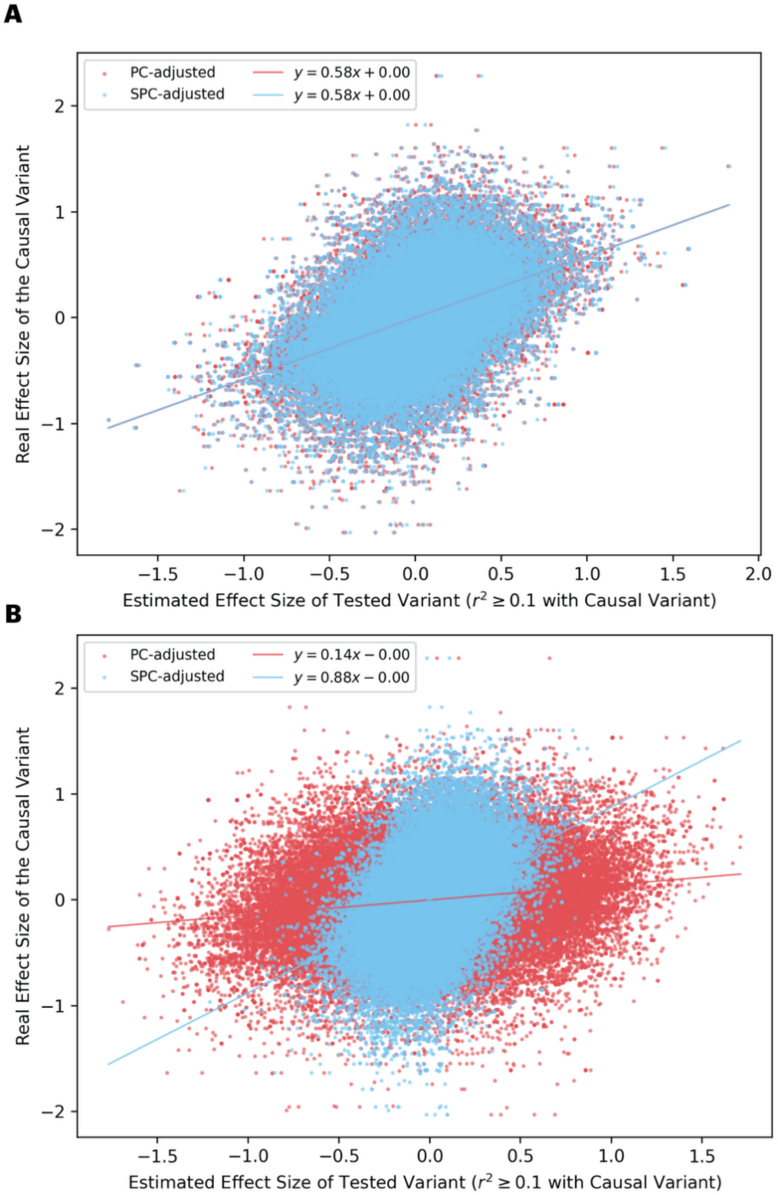


**Supplemental Figure 4 –** Effects of the confounding factors of environmental noise on the calibration of models. The x-axes display the estimated effect size of tagging variants (LD with causal variant > 0.1). The y-axes show the true effect size assigned to the tagged causal variant. The slope of regression lines measured the overall concordance between the true effect sizes and the estimated ones, both in the absence of confounding using random noise in the polygenic heritable phenotype (**a**), and in its presence, using hybrid smooth phenotype (**b**).

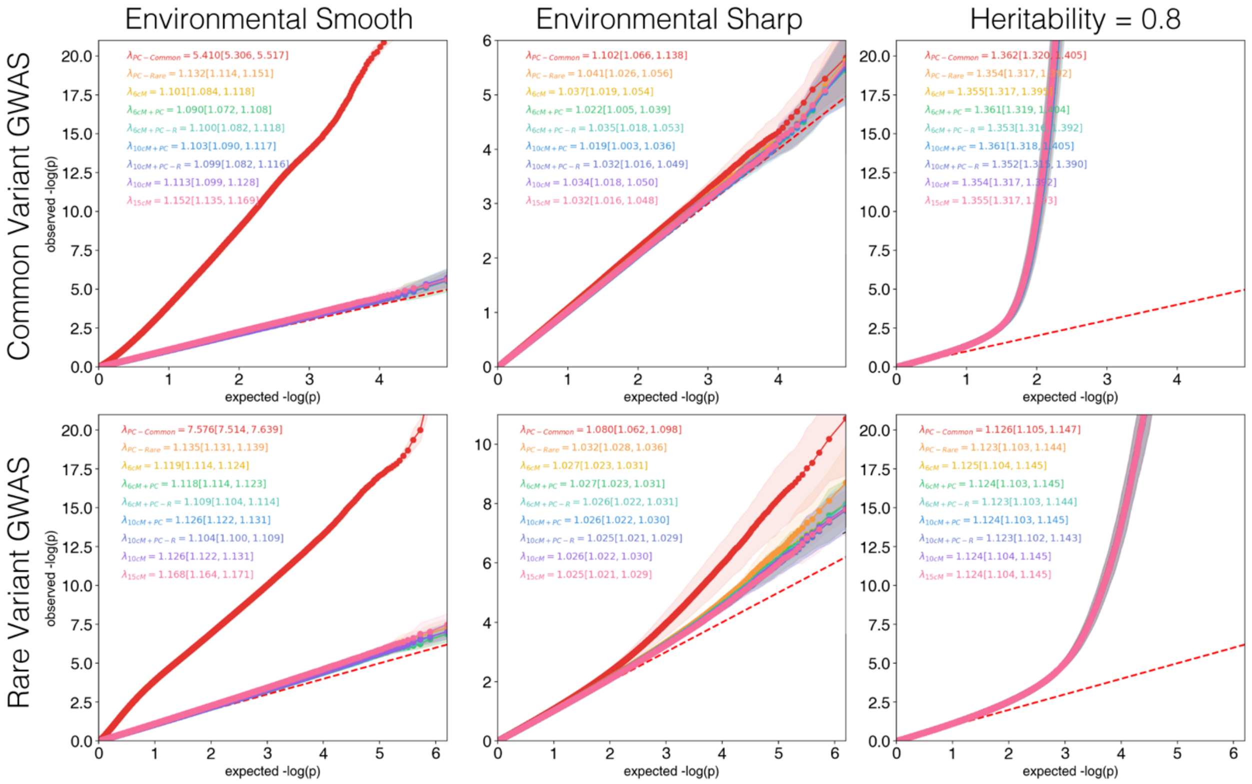


**Supplemental Figure 5 -** Genomic inflation of the results of GWAS analysis of simulated phenotypes in a simulated 5 by 5 grid of demes with 50,000 samples. 3 phenotypes shown here are, from left to right, environmental smooth, environmental sharp, and polygenic (high heritability; $h^2$=0.8) phenotypes. The top panels show the inflation in the results of common variants GWAS, and the bottom panel show the results of rare variants (10<MAC<500) GWAS. The text on each panel displays the genomic inflation factor, along with its credible interval, for 9 tested models. From top to bottom, these models are: PCs of common variants, PCs of rare variants, SPCs, SPCs and common variant PCs combined, SPCs and rare variant PCs combined, SPCs (calculated using a 10cM threshold) combined with common variant PCs, SPCs (calculated using a 10cM threshold) combined with rare variant PCs, SPCs (calculated using a 10cM threshold), and SPCs (calculated using a 15cM threshold).

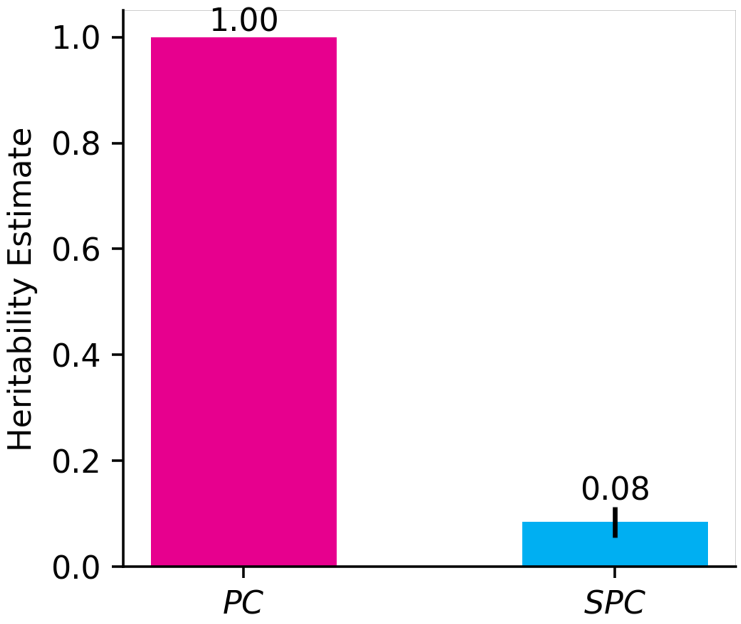


**Supplemental Figure 6:** narrow-sense heritability analysis aims to measure the proportion of heritability in a phenotype that is derived from genetic factors. Our analysis of a simulated 'environmental smooth' phenotype with no genetic effects using PCs or SPCs as covariates illustrates how SPCs are better suited for such analysis in phenotypes strongly affected by environmental factors. Adjusting for PCs results in a heritability estimate of 1.00 while adjusting for SPCs yields a much more realistic estimate of 0.08.

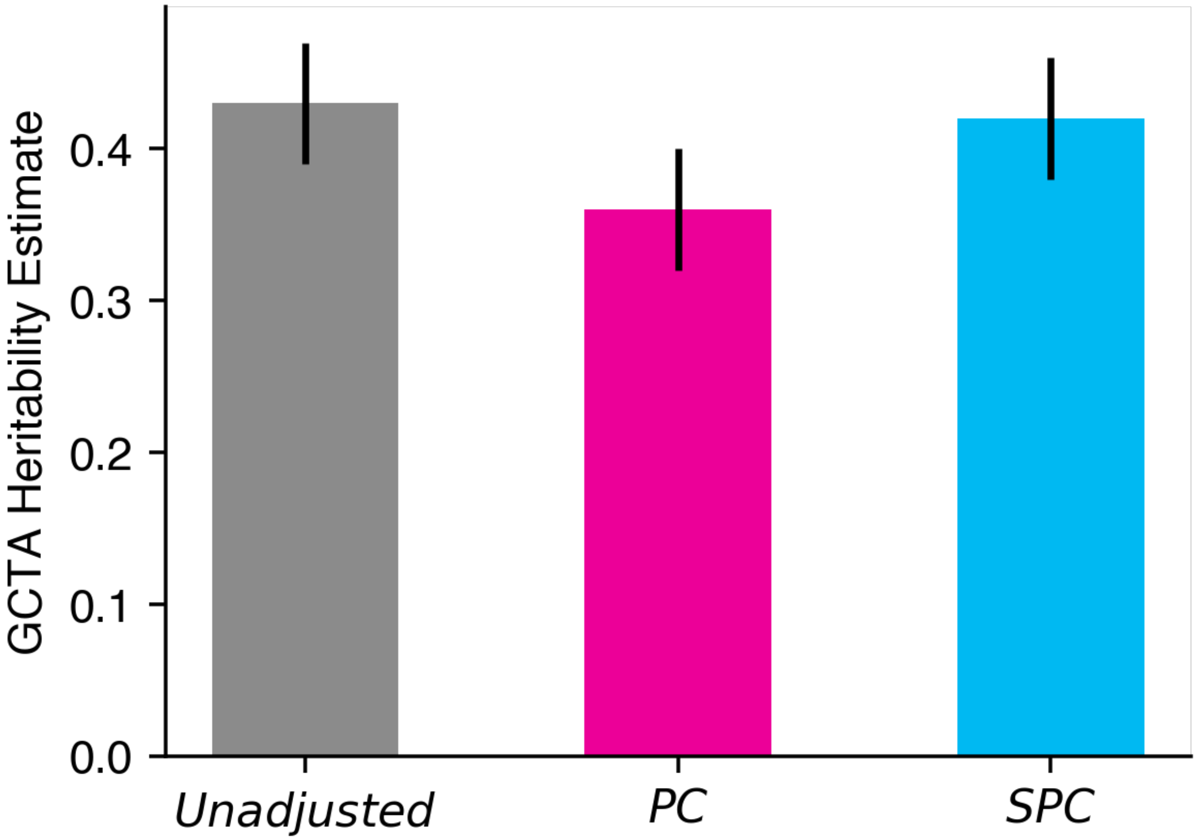


**Supplemental Figure 7:** narrow-sense heritability analysis aims to measure the proportion of heritability in a phenotype that is derived from genetic factors. Our analysis of a simulated phenotype with high heritability (h=0.8) and lower polygenicity (causal variants windows size 100,000 vs. 10,000 in the main experiments) aims to measure overcorrection of heritability estimates by each model in lower polygenicity settings. Heritability estimates were calculated using GREML method in the GCTA software package.

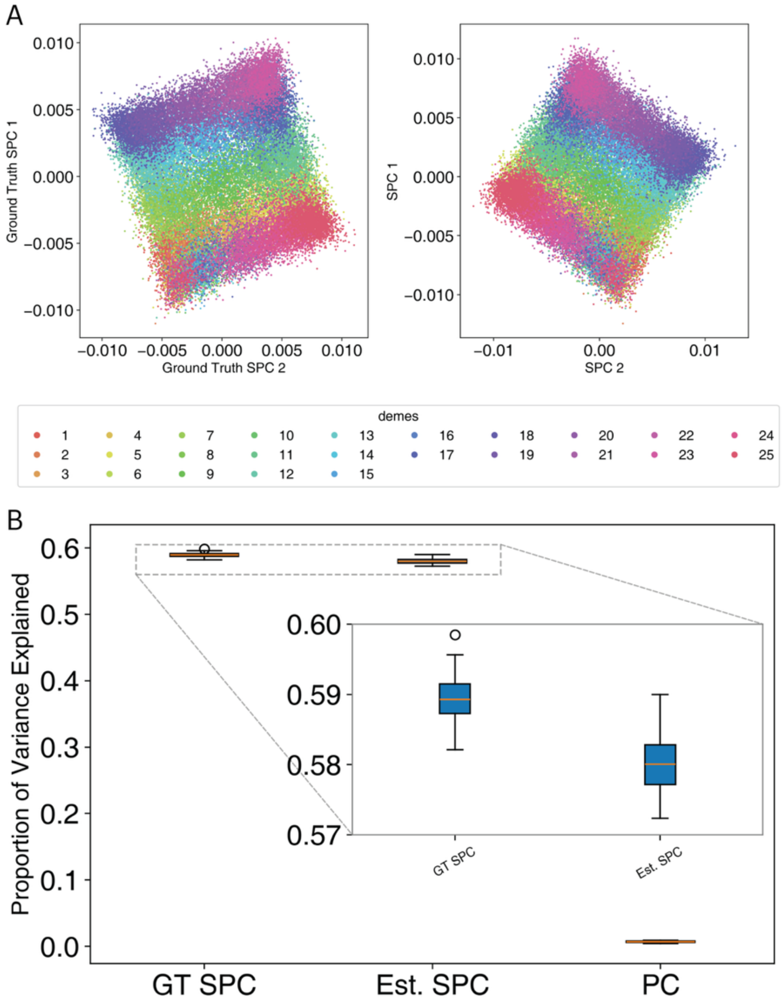


**Supplemental Figure 8:** Performance comparison between estimated and ground truth SPCs. Estimated SPCs were derived from unphased common variant data through phasing and IBD estimation steps, processes that can introduce estimation errors. Ground truth SPCs were calculated directly from the reference simulated ARG structure. A) First two dimensions from ground truth SPCs (left) and estimated SPCs (right). Each dot represents a simulated diploid individual, colored by deme of origin. B) Proportion of variance explained in the hybrid smooth phenotype by the first 25 components from Ground Truth SPCs (GT SPC), Estimated SPCs (Est. SPC), and standard PCs of common variants.

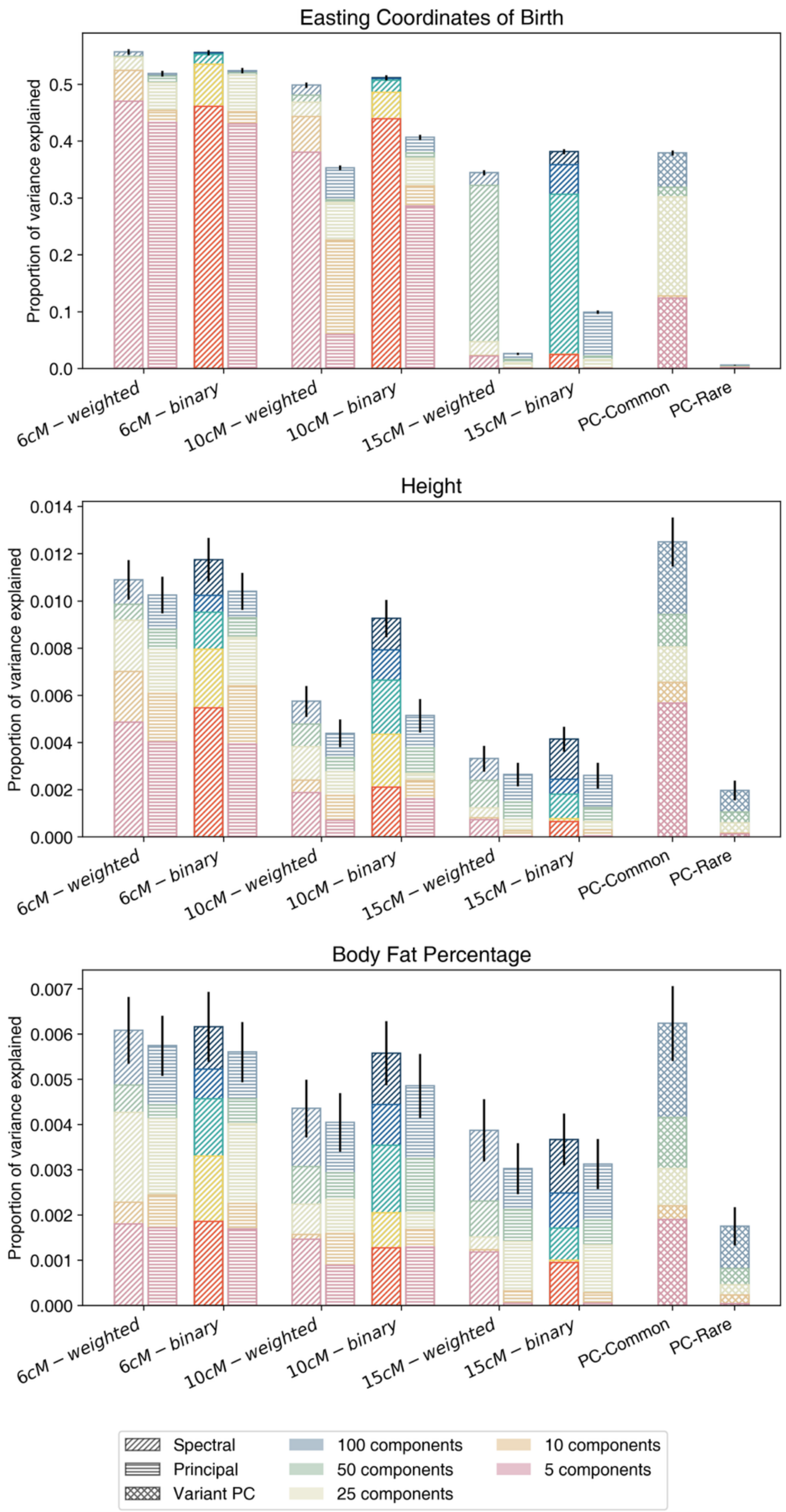


**Supplemental Figure 9 –** Proportion of variance in eastings, BFP, and height explained by principal components in the UK Biobank. Exploring the effects of minimum IBD sharing parameter, graph weighting scheme, and component calculation technique. Principal components of common and rare variants are displayed on the rightmost side of the figure, labeled as PC-

Common, and PC-Rare, respectively. For other components, labels on the x-axis indicate the data type used to generate the components. Spectral and principal components of each data type were calculated, indicated by diagonal and horizontal shading lines, respectively. Components calculated from weighted graphs, where weights are assigned based on total IBD sharing between pairs are subscripted as weighted. Components calculated from binary graphs are subscripted as binary. SPCs, spectral components calculated from unweighted graphs, are highlighted in brighter colors.

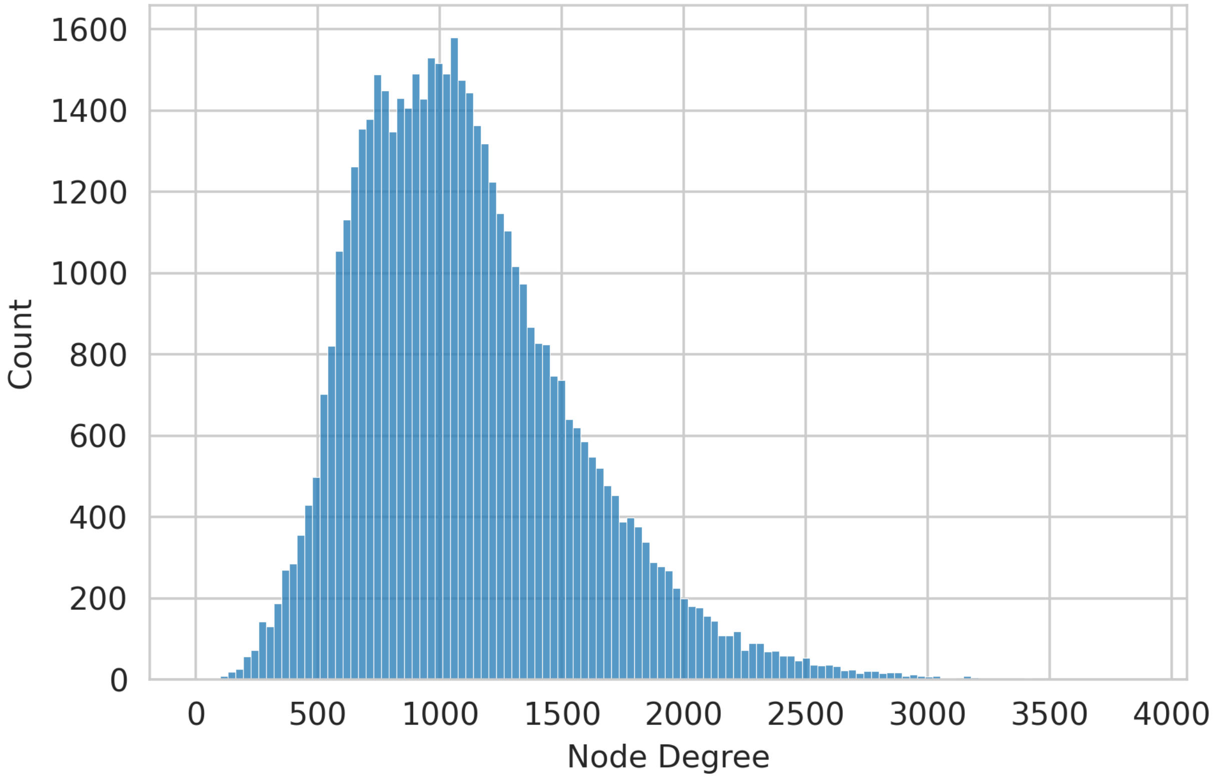


**Supplemental Figure 10 –** Degree distribution in a randomly selected sample (N=50,000) of the global IBD sharing graph in the UK Biobank. The node degrees are calculated as the total number of nodes connected to each focal node in the subsample.

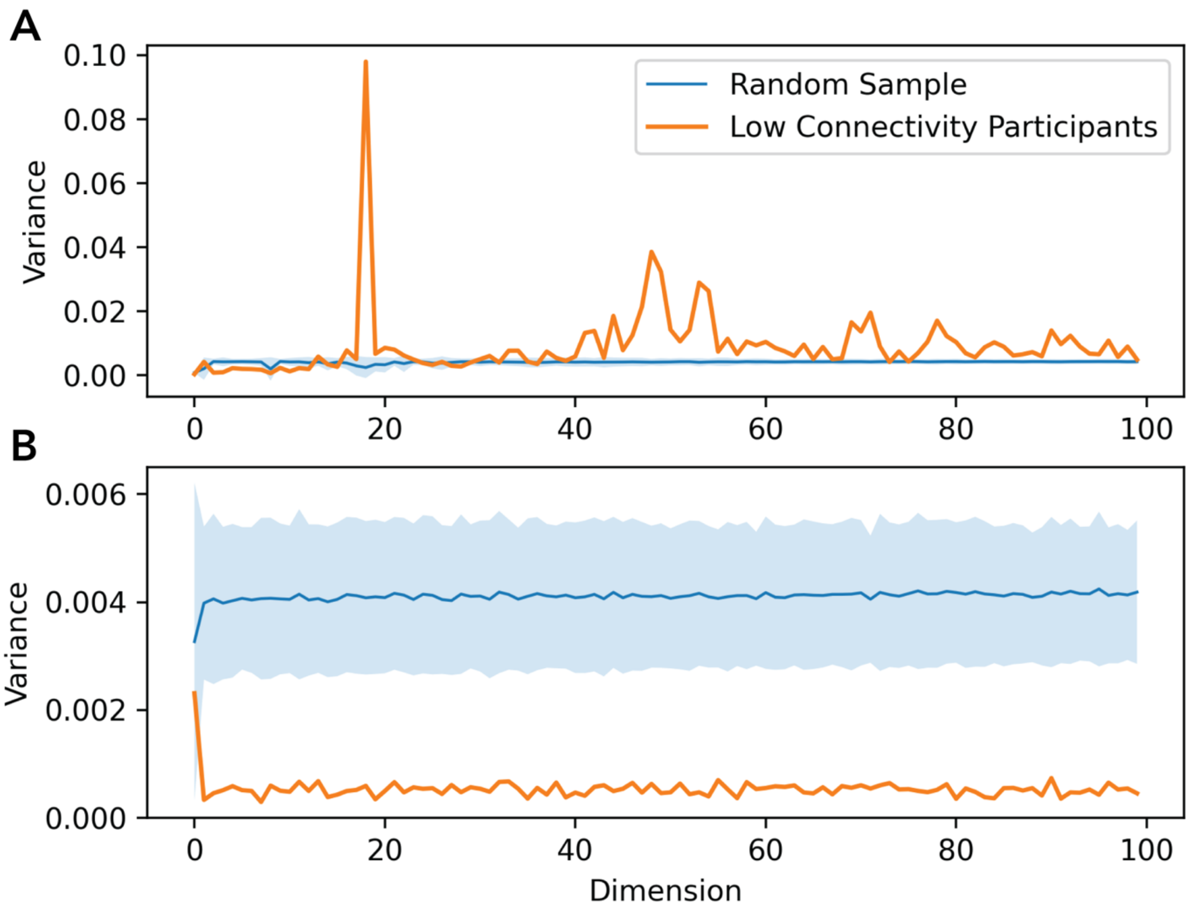


**Supplemental Figure 11 –** Per-dimension variance of component values calculated for the 20 nodes with the lowest connectivity in the graph, versus that of a random sample of the same size in (A) SPC latent space, and (B) PC latent space. The PC representations utilized were calculated using the same IBD sharing graph as SPCs. Credible area calculated from 1,000 repetitions of the experiment.

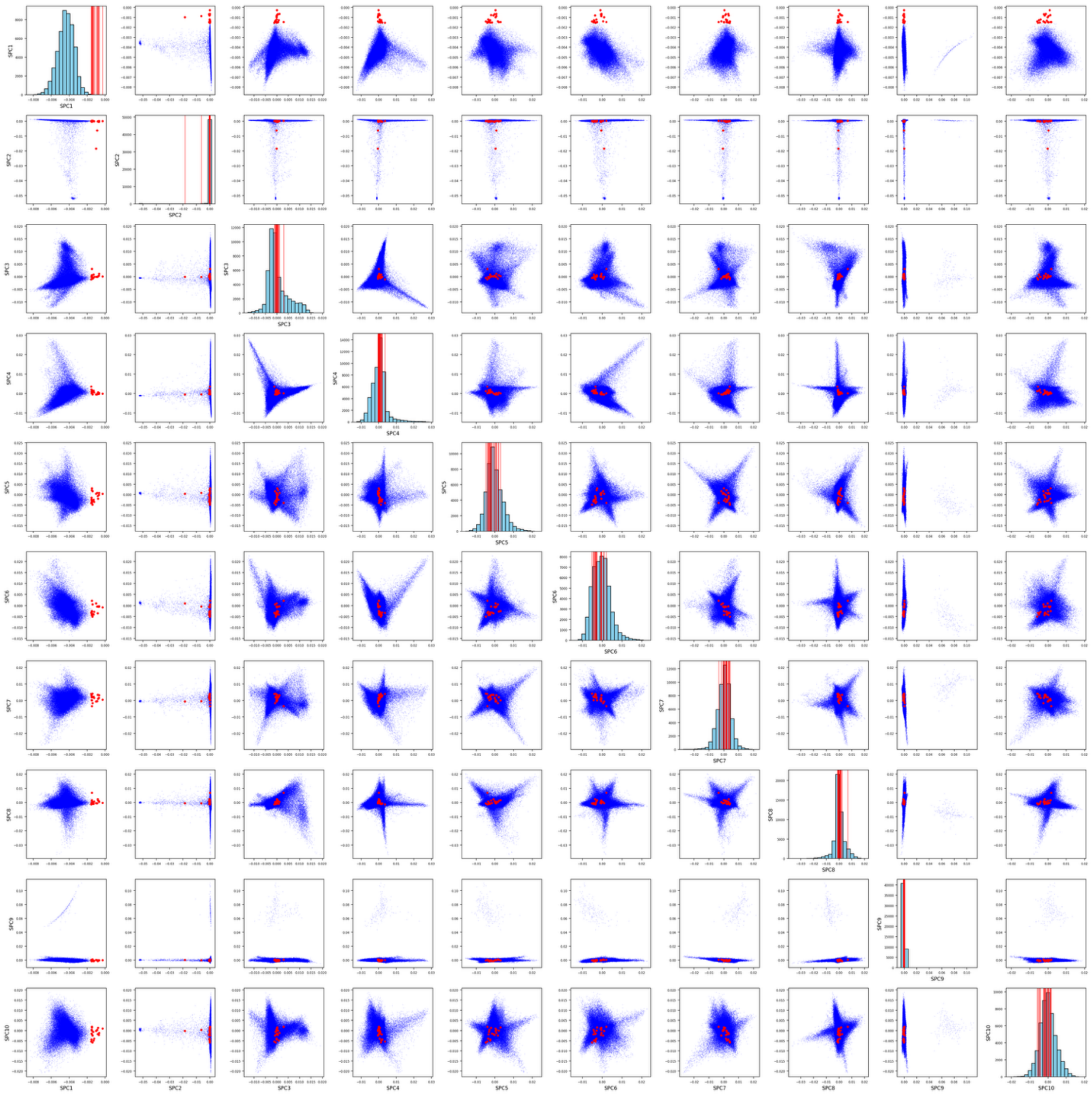

**Supplemental Figure 12 –** Scatter plot and histogram plots of the first 10 SPCs dimensions against other dimensions. Nodes representing the bottom 20 individuals in terms of connectivity are colored in red. The diagonal cells display the histogram of the assigned dimension. Vertical lines indicate the values of the latent representation of the 20 individuals in each dimension's histogram.

| Phenotype | Covariate Count | SPC (6cM) | PCs | Rare variant PCs | IBD PC (Weighted; 6cM) | Weighted SPC (6cM) | IBD PC (Binary;6cM) | SPC (10cM) | SPC (15cM) |
|---|---|---|---|---|---|---|---|---|---|
| Hybrid Smooth | 5 | 0.5800-0.0042 | 0.0008-0.0005 | 0.5798-0.0041 | 0.5599-0.0040 | 0.5871-0.0040 | 0.5678-0.0042 | 0.5796-0.0041 | 0.5742-0.0039 |
| | 10 | 0.5802-0.0042 | 0.0025-0.0008 | 0.5800-0.0041 | 0.5605-0.0040 | 0.5871-0.0040 | 0.5679-0.0042 | 0.5797-0.0041 | 0.5743-0.0039 |
| | 25 | 0.5805-0.0042 | 0.0068-0.0013 | 0.5804-0.0041 | 0.5615-0.0040 | 0.5875-0.0040 | 0.5683-0.0042 | 0.5808-0.0042 | 0.5753-0.0039 |
| | 50 | 0.5812-0.0042 | 0.0125-0.0017 | 0.5810-0.0041 | 0.5630-0.0040 | 0.5884-0.0040 | 0.5691-0.0042 | 0.5813-0.0042 | 0.5760-0.0039 |
| | 100 | 0.5822-0.0042 | 0.0310-0.0024 | 0.5821-0.0040 | 0.5652-0.0039 | 0.5897-0.0040 | 0.5702-0.0041 | 0.5822-0.0042 | 0.5767-0.0039 |
| Hybrid Sharp | 5 | 0.0111-0.0064 | 0.0006-0.0005 | 0.0091-0.0057 | 0.0085-0.0058 | 0.0114-0.0066 | 0.0108-0.0063 | 0.0111-0.0063 | 0.0111-0.0060 |
| | 10 | 0.0165-0.0080 | 0.0009-0.0005 | 0.0102-0.0057 | 0.0090-0.0058 | 0.0148-0.0069 | 0.0155-0.0076 | 0.0169-0.0082 | 0.0169-0.0083 |
| | 25 | 0.0175-0.0080 | 0.0020-0.0008 | 0.0145-0.0066 | 0.0130-0.0062 | 0.0178-0.0083 | 0.0162-0.0077 | 0.0188-0.0081 | 0.0196-0.0085 |
| | 50 | 0.0188-0.0082 | 0.0039-0.0012 | 0.0166-0.0073 | 0.0158-0.0070 | 0.0193-0.0084 | 0.0173-0.0078 | 0.0204-0.0082 | 0.0211-0.0084 |
| | 100 | 0.0207-0.0081 | 0.0075-0.0015 | 0.0189-0.0075 | 0.0187-0.0074 | 0.0224-0.0084 | 0.0193-0.0079 | 0.0227-0.0082 | 0.0235-0.0085 |
| heritable (h=0.1) | 5 | 0.0001-0.0000 | 0.0002-0.0001 | 0.0001-0.0000 | 0.0001-0.0001 | 0.0001-0.0000 | 0.0001-0.0000 | 0.0001-0.0001 | 0.0001-0.0000 |
| | 10 | 0.0002-0.0001 | 0.0003-0.0002 | 0.0002-0.0001 | 0.0002-0.0001 | 0.0002-0.0001 | 0.0002-0.0001 | 0.0002-0.0001 | 0.0002-0.0001 |
| | 25 | 0.0005-0.0001 | 0.0009-0.0004 | 0.0005-0.0002 | 0.0005-0.0002 | 0.0005-0.0001 | 0.0005-0.0001 | 0.0006-0.0002 | 0.0005-0.0001 |
| | 50 | 0.0011-0.0001 | 0.0019-0.0005 | 0.0011-0.0002 | 0.0011-0.0002 | 0.0010-0.0002 | 0.0011-0.0002 | 0.0011-0.0002 | 0.0010-0.0002 |
| | 100 | 0.0022-0.0002 | 0.0037-0.0006 | 0.0022-0.0003 | 0.0022-0.0004 | 0.0022-0.0004 | 0.0022-0.0003 | 0.0022-0.0003 | 0.0021-0.0002 |
| heritable (h=0.3) | 5 | 0.0002-0.0001 | 0.0004-0.0003 | 0.0002-0.0001 | 0.0002-0.0001 | 0.0002-0.0001 | 0.0002-0.0001 | 0.0002-0.0001 | 0.0002-0.0001 |
| | 10 | 0.0003-0.0002 | 0.0007-0.0003 | 0.0003-0.0002 | 0.0003-0.0002 | 0.0003-0.0001 | 0.0003-0.0002 | 0.0003-0.0002 | 0.0003-0.0002 |
| | 25 | 0.0007-0.0003 | 0.0017-0.0007 | 0.0008-0.0003 | 0.0007-0.0002 | 0.0007-0.0002 | 0.0007-0.0002 | 0.0008-0.0002 | 0.0007-0.0002 |
| | 50 | 0.0013-0.0002 | 0.0036-0.0009 | 0.0014-0.0004 | 0.0014-0.0003 | 0.0014-0.0003 | 0.0012-0.0003 | 0.0014-0.0003 | 0.0014-0.0003 |
| | 100 | 0.0025-0.0003 | 0.0071-0.0009 | 0.0027-0.0005 | 0.0026-0.0004 | 0.0026-0.0003 | 0.0025-0.0003 | 0.0026-0.0004 | 0.0027-0.0003 |
| heritable (h=0.8) | 5 | 0.0003-0.0001 | 0.0012-0.0012 | 0.0003-0.0001 | 0.0003-0.0001 | 0.0003-0.0001 | 0.0003-0.0001 | 0.0003-0.0001 | 0.0003-0.0001 |
| | 10 | 0.0005-0.0002 | 0.0017-0.0013 | 0.0005-0.0002 | 0.0005-0.0002 | 0.0005-0.0002 | 0.0005-0.0002 | 0.0005-0.0001 | 0.0005-0.0002 |
| | 25 | 0.0010-0.0003 | 0.0040-0.0017 | 0.0011-0.0003 | 0.0010-0.0003 | 0.0010-0.0003 | 0.0010-0.0003 | 0.0010-0.0003 | 0.0009-0.0003 |
| | 50 | 0.0019-0.0005 | 0.0080-0.0026 | 0.0021-0.0004 | 0.0019-0.0005 | 0.0019-0.0004 | 0.0018-0.0004 | 0.0018-0.0003 | 0.0017-0.0004 |
| | 100 | 0.0036-0.0003 | 0.0149-0.0038 | 0.0039-0.0006 | 0.0037-0.0006 | 0.0038-0.0007 | 0.0035-0.0004 | 0.0035-0.0004 | 0.0034-0.0004 |
| Environmental Smooth | 5 | 0.8293-0.0006 | 0.0007-0.0001 | 0.8288-0.0007 | 0.8001-0.0006 | 0.8391-0.0006 | 0.8118-0.0007 | 0.8287-0.0006 | 0.8208-0.0006 |
| | 10 | 0.8294-0.0006 | 0.0025-0.0001 | 0.8290-0.0007 | 0.8009-0.0006 | 0.8391-0.0006 | 0.8119-0.0007 | 0.8287-0.0006 | 0.8208-0.0006 |
| | 25 | 0.8298-0.0006 | 0.0074-0.0002 | 0.8293-0.0007 | 0.8019-0.0006 | 0.8393-0.0006 | 0.8122-0.0007 | 0.8299-0.0006 | 0.8221-0.0007 |
| | 50 | 0.8303-0.0006 | 0.0139-0.0003 | 0.8297-0.0007 | 0.8036-0.0006 | 0.8401-0.0005 | 0.8129-0.0007 | 0.8304-0.0006 | 0.8227-0.0007 |
| | 100 | 0.8309-0.0006 | 0.0366-0.0005 | 0.8303-0.0007 | 0.8058-0.0006 | 0.8412-0.0005 | 0.8137-0.0007 | 0.8308-0.0006 | 0.8231-0.0007 |
| Environmental Sharp | 5 | 0.0158-0.0093 | 0.0001-0.0001 | 0.0129-0.0083 | 0.0121-0.0084 | 0.0163-0.0096 | 0.0154-0.0092 | 0.0159-0.0091 | 0.0158-0.0085 |
| | 10 | 0.0235-0.0121 | 0.0003-0.0001 | 0.0144-0.0084 | 0.0127-0.0085 | 0.0210-0.0102 | 0.0222-0.0115 | 0.0242-0.0123 | 0.0242-0.0123 |
| | 25 | 0.0247-0.0120 | 0.0007-0.0002 | 0.0205-0.0098 | 0.0184-0.0092 | 0.0252-0.0125 | 0.0230-0.0116 | 0.0267-0.0122 | 0.0279-0.0125 |
| | 50 | 0.0262-0.0120 | 0.0014-0.0004 | 0.0230-0.0108 | 0.0220-0.0104 | 0.0270-0.0126 | 0.0241-0.0117 | 0.0286-0.0123 | 0.0298-0.0125 |
| | 100 | 0.0280-0.0119 | 0.0028-0.0005 | 0.0254-0.0110 | 0.0253-0.0111 | 0.0306-0.0127 | 0.0262-0.0118 | 0.0310-0.0124 | 0.0324-0.0126 |

**Supplemental Table 1 -** Proportion of variance explained in simulated phenotypes by each covariate model. Variance of each phenotype is modeled using the first 5, 10, 25, 50, 100 components from each model. In addition to the PCs of common and rare variants, 3 different SPC sets (using minimum IBD sharing cutoffs of 6cM,10cM, and 15cM), PCs of IBD sharing matrix (both weighted using the total IBD sharing and unweighted or binary at 6cM cutoff), and spectral components of weighted IBD sharing matrix (at 6cM cutoff) are displayed. The first number in each cell is the mean calculated from 2000 repetitions, while the second number is the standard deviation.

| Phenotype | PC-adjusted Heritability | SPC-adjusted Heritability |
|---|---|---|
| **heritable (h=0.1)** | 0.047 - 0.004 | 0.046 - 0.004 |
| **heritable (h=0.3)** | 0.119 - 0.005 | 0.118 - 0.005 |
| **heritable (h=0.8)** | 0.317 - 0.006 | 0.317 - 0.006 |
| **Environmental Sharp** | 0.11 - 0.04 | 0.02 - 0.04 |
| **Environmental Smooth** | 1.00 - 0.001 | 0.08 - 0.03 |
| **Hybrid Smooth** | 0.64 – 0.005 | 0.30 – 0.006 |
| **Hybrid Sharp** | 0.14 – 0.005 | 0.13 – 0.005 |

**Supplemental Table 2 –** Estimates of heritability for each phenotype after adjustment using PCs and SPCs. Heritability estimates were calculated using the GREML functionality implemented in the GCTA software package. estimated means and standard deviations are reported in each cell.

#### Supplemental methods

##### Performance against alternative approaches

We measured the efficacy of alternative strategies in accounting for recent population structure by calculating the total proportion of variance explained in each phenotype by them.

##### Principal Components of rare variants

We calculated PCs of rare variants using two thresholds for rarity. First, all variants with MAF<0.01. Second, variants with minor allele counts of 2-4 (Zaidi & Mathieson, 2020). We are only reporting the results of the first definition as it performed better in all scenarios in our simulations. We found the performance of rare variant PCs to be inconsistent compared to common variant PCs. Rare variant PCs explained a higher proportion of variance compared to PCs in the environmental phenotypes (**Supplemental Figure 1**). Subsequently, the genomic inflation factor in the GWAS of those phenotypes was lower when adjusted using rare PCs, both in the GWAS of common and rare variants (**Supplemental Figure 5**). However, they explained a lower proportion of variance in the polygenic phenotype, although that did not translate to any significant difference in the inflation of GWAS results. Rare variants lowered the heritability estimate of the environmental smooth phenotype, from 1.00 to 0.45, compared to PCs. However, they underperformed in comparison to SPCs across all simulated scenarios. Their performance in the analysis of the polygenic phenotype was not significantly different compared to SPCs. The performance of SPCs further deteriorated in the analysis of real data. Using rare variant data extracted from WES data for 50,000 participants in the UK Biobanks, we found that PCs of common variants have a higher PVE for both eastings and height compared to PCs of rare variants (**Supplemental Figure 2**).

##### Alternative IBD-based covariates

We calculated 11 alternative IBD-based covariates. These alternative covariates varied from SPCs in three aspects. First, while SPCs are *spectral components* of the IBD relatedness, one can also calculate *principal components* of IBD relatedness. Second, the IBD relatedness can either be expresses as a binary or weighted relationship. Unlike the unweighted binary relationship, in the weighted IBD relatedness graphs, higher weights are assigned to edges connecting pairs of samples that share more than the minimum threshold of sharing. These weights are calculated by aggregating the lengths of IBD segments shared between each pair of individuals. Finally, the minimum threshold of IBD sharing itself is treated as a parameter. We heuristically chose three different minimum thresholds of 6cM, 10cM, and 15cM. We used proportion of explained variance as the comparison criteria (**Supplemental Figures 1 and 9**).

Covariates generated using binary similarity matrix outperformed those derived from the weighted matrix when adjusting for the environmental phenotypes in simulation, and for all three phenotypes in the UK Biobank, especially as the minimum IBD threshold was increased. Simultaneously, the latter group showed a higher correlation with PCs of common and rare variants, suggesting a higher level of overlap in the signals represented by the covariates.

Spectral components (including SPCs) outperformed principal components. The advantage of spectral components over principal components was small but maintained across both environmental scenarios. This difference was statistically significant for the environmentally smooth phenotype, yet not significant for the sharp phenotype, most likely due to its nonlinear structure. The gap between SPCs and principal components increased in the analysis of easting, BFP, and height in UK Biobank, across all minimum length thresholds. The first 5 SPCs calculated using a binary network with a minimum threshold of 10cM had a higher PVE for eastings compared to the first 100 principal components calculated using the same network (**Supplemental Figure 9**). There was a significant difference between spectral components and principal components calculated from the weighted matrix, especially as the minimum IBD threshold is increased. SPCs have 67% higher performance at 6cM, 84% at 10cM, and 92% at 15cM when predictive the sharp phenotype.

Increasing the minimum threshold of IBD sharing used to generate the relatedness network had negligible effect on PVE in simulated phenotypes. Consequently, it did not significantly change the inflation of p-values in the GWAS of the simulated phenotypes. However, in our analysis of PVE of height, BFP, and eastings in UK Biobank, increasing this threshold significantly lowered PVE, to the point where the PVE of SPCs generated using the 15cM network were lower than those of PCs in the analysis of eastings.

**Graph structure captured by SPCs**

Spectral components can recover non-linear properties in a graph. Here we will describe what that entails for the characteristics of the population structure they extract from IBD relatedness graphs. SPCs attribute a set of numerical values to each vertex based on its projections on the set of principal axes of variation in the graph. The level of detail represented by each axis depends on the corresponding eigenvalue association with it. Eigenvectors associated with the smaller eigenvalues will assign similar numbers to neighbors on the graph, whereas the eigenvector associated with the larger eigenvalues will assign varying numbers to neighboring nodes. Thus, ignoring the axes with eigenvalues equal or very close to zero, the first axes, ordered by the magnitude of their eigenvalues attached to them, will capture the most polarizing aspects of variations in the relatedness in the graph with lowest level of granularity. Components with zero, or close to zero eigenvalues incorporate a clustering of vertices into groups of recent genetic ancestry where participants from the major ancestry group $\boldsymbol{a}$ are represented by nonzero values with the mean $1/\sqrt{n_a}$, while other samples, with low, or no connections to this ancestry group, are represented by values closer to zero in that dimension. This dependency on connection, and not balance in representation, is among the distinctions between SPCs and PCs (Lee et al., 2010). The number of highly distinctive features is thus derived from the number of distinct IBD families present in the dataset with heavy connections. An extreme example happens if the dataset is comprised of two heavily connected familial groups (founder populations) with little or no connections to each other. Such groups would be represented by their own dimension, even if they are not very well represented in the ascertainment. The SPCs also include dimensions that represent cross-family similarities. These dimensions have higher than zero eigenvalues associated with them. Thus, in the absence of strong clustering (i.e. a homogeneous cohort), the SPCs can still represent overlapping groups of individuals with recent genetic similarities.
